# De novo variants in the *PSMC3* proteasome AAA-ATPase subunit gene cause neurodevelopmental disorders associated with type I interferonopathies

**DOI:** 10.1101/2021.12.07.21266342

**Authors:** Frédéric Ebstein, Sébastien Küry, Victoria Most, Cory Rosenfelt, Marie-Pier Scott- Boyer, Geeske M. van Woerden, Thomas Besnard, Jonas Johannes Papendorf, Maja Studencka-Turski, Tianyun Wang, Tzung-Chien Hsieh, Richard Golnik, Dustin Baldridge, Cara Forster, Charlotte de Konink, Selina M.W. Teurlings, Virginie Vignard, Richard H. van Jaarsveld, Lesley Ades, Benjamin Cogné, Cyril Mignot, Wallid Deb, Marjolijn C.J. Jongmans, F. Sessions Cole, Marie-José H. van den Boogaard, Jennifer A. Wambach, Daniel J. Wegner, Sandra Yang, Vickie Hannig, Jennifer Ann Brault, Neda Zadeh, Bruce Bennetts, Boris Keren, Anne-Claire Gélineau, Zöe Powis, Meghan Towne, Kristine Bachman, Andrea Seeley, Anita E. Beck, Jennifer Morrison, Rachel Westman, Kelly Averill, Theresa Brunet, Judith Haasters, Melissa T. Carter, Matthew Osmond, Patricia G. Wheeler, Francesca Forzano, Shehla Mohammed, Yannis Trakadis, Andrea Accogli, Rachel Harrison, Deciphering Developmental Disorders Study, Care4Rare Canada Consortium, Sophie Rondeau, Geneviève Baujat, Giulia Barcia, René Günther Feichtinger, Johannes Adalbert Mayr, Martin Preisel, Frédéric Laumonnier, Alexej Knaus, Bertrand Isidor, Peter Krawitz, Uwe Völker, Elke Hammer, Arnaud Droit, Evan E. Eichler, Ype Elgersma, Peter W. Hildebrand, François Bolduc, Elke Krüger, Stéphane Bézieau

## Abstract

A critical step in preserving protein homeostasis by the ubiquitin-proteasome system (UPS) is the recognition, binding, unfolding, and translocation of protein substrates by AAA-ATPase proteasome subunits for degradation by 26S proteasomes. Here, we identified fourteen different *de novo* missense variants in the *PSMC3* gene encoding the AAA-ATPase proteasome subunit Rpt5 in twenty-two unrelated heterozygous subjects with an autosomal dominant form of neurodevelopmental delay and intellectual disability. Indeed, depletion of *PSMC3* impaired reversal learning capabilities in a *Drosophila* model. The *PSMC3* variants cause proteasome dysfunction in patient-derived cells by disruption of substrate translocation, proteotoxic stress and proteostatic imbalances, as well as alterations in proteins controlling developmental and innate immune programs. Molecular analysis confirmed the induction of cellular stress responses and dysregulated mitophagy along with an elevated type I interferon (IFN) signature. Our data define *PSMC3* variants as the genetic cause of proteotoxic stress alerting the innate immune system to mount a type I IFN response and link neurodevelopmental syndromes to interferonopathies.

## Introduction

Proteasomes are large multi-protein complexes whose structure is adapted to the function of protein degradation [1, 2]. Together with the autophagosomal-lysosomal system, it maintains protein homeostasis by counterbalancing the synthesis of new proteins by the translational machinery [3–6]. The proteasome is part of the ubiquitin-proteasome system (UPS) which counts numerous enzymes acting upstream of the proteasome [7, 8]. Aged or unstructured proteins are preliminary ubiquitin-tagged by the ubiquitination machinery through a cascade of enzymatic reactions [9, 10]. Ubiquitin modification is a signal recognized by the 26S proteasome which then takes in charge the hydrolysis of the substrate.

The 26S proteasome consists of two parts, the 20S core proteolytic particle and the 19S regulatory particle which caps the 20S particle at one and/or both ends [11, 12]. Briefly, polyubiquitinated proteins are recognized by the 19S regulatory particle where they are unfolded and deubiquitinated before their ATP-dependent translocation into the 20S core 1 and β2 catalytic subunits that exhibit chymotrypsin-, trypsin and caspase-like activities, respectively [15, 16]. Depending on the nature of the catalytic subunits, two major forms of proteasomes exist, namely the standard ones containing the β1, β2 and β5 catalytic standard subunits and the immunoproteasomes harboring the β1i, β2i and β5i inducible subunits [17]. While standard proteasomes are expressed in virtually all types of tissues, the expression of the inducible subunits is restricted to immune cells and non-immune cells exposed to type I and/or II interferons (IFN) [18, 19].

In more details, the 19S regulatory particle comprises two parts: a base and a lid. In the base, four regulatory particle non-ATPase subunits (*i.e.* Rpn1, Rpn2, Rpn10 and Rpn13) ensure recognition and capture of ubiquitin-modified substrates [20], whereas six AAA (<u>A</u>TPase <u>A</u>ssociated with various cellular <u>A</u>ctivities)-ATPase subunits, namely PSMC1-6 (also referred to as Rpt1-6) use the energy provided by ATP hydrolysis to unfold and translocate the substrate into the 20S proteolytic core particle by gate opening [21]. The lid contains eight non-ATPase scaffolding subunits (Rpn3, Rpn5-9, Rpn12, and Rpn15) [22, 23], and the deubiquitinating enzyme (DUB) Rpn11 [24].

Pathogenic variants in proteasome subunit genes cause rare proteasomopathies with a broad spectrum of symptoms [25, 26]. So far, with the exception of the PSMB1 (i.e. β subunit [27], all pathogenic variants related to the 20S core particle have been shown to provoke immune dysregulation. Indeed, several genes encoding β-subunits (*PSMB4*, *PSMB8*, *PSMB9*, *PSMB10*), α-subunits (*PSMA3*) or assembly chaperone genes (*POMP, PSMG2)* of the 20S proteasome complex have been involved in autosomal recessive proteasome-associated autoinflammatory syndromes (PRAAS) [28–35]. By contrast, the only Mendelian disorder involving a gene of the 19S regulatory particle, the Stankiewicz-Isidor syndrome (MIM: 617516) caused by truncating variants of *PSMD12* (also referred to as Rpn5) is a neurodevelopmental polymalformative syndrome [36], recently associated with a mild interferonopathy (Isidor et al., in press).

In this work, we identify a series of fourteen dominant *de novo* variants in the *PSMC3* gene coding for the AAA-ATPase PSMC3/Rpt5. Rare missense variants were detected in twenty-two individuals presenting with neurodevelopmental delay (NDD) and/or intellectual disability (ID) together with various congenital malformations. Together, our data highlight interferonopathy as a potential contributor to the pathogenesis of NDD/ID in subjects carrying loss-of-function variants in subunits of the 19S proteasome regulatory particle.

## Materials & Methods

### Genetic studies and ethics statement

This study was approved by the CHU de Nantes ethics committee (Research Programme “Génétique Médicale DC-2011-1399). Written informed consent was obtained from all study participants, including probands and healthy parents. All affected individuals were initially referred for unexplained developmental delay (DD) and/or intellectual disability (ID) together with various congenital malformations. They underwent extensive clinical examination by at least one expert clinical geneticist. Routine genetic testing was performed whenever clinically relevant, including copy number variation (CNV) analysis by high-resolution array-based comparative genomic hybridization (aCGH). As these tests provided no diagnosis, trio-based whole exome sequencing (WES) was performed on a diagnostic or research setting whenever parental samples were available.

### Patient samples

Peripheral blood mononuclear cells (PBMC) used in this paper were isolated from blood draws from patients and related healthy controls (father and/or mother of the proband). Briefly, PBMC were isolated by PBMC spin medium gradient centrifugations (pluriSelect), washed three times with PBS, frozen in FBS with 10% DMSO and stored in liquid nitrogen for further use. In some experiments, collected PBMC were expanded in U-bottom 96-well plates together with feeder cells using RPMI 1640 supplemented with 10% human AB serum (both purchased from PAN-Biotech GmbH) in the presence of 150 U/ml IL-2 (Miltenyi Biotec) and 1 µg/µl L-PHA (Sigma) following the procedure of Fonteneau et al. [37]. After 3-4 weeks of culture, resting T cells were washed and frozen as dry pellets for further use.

### SDS-PAGE and western-blot analysis

Cell pellets from resting T cell isolated from patients and related controls were lysed in equal amounts of standard RIPA buffer (50 mM Tris pH 7.5, 150 mM NaCl, 2 mM EDTA, 1 mM N-ethylmaleimide, 10 µM MG-132, 1% NP40, 0.1% SDS) and separated by 10 or 12.5% SDS-PAGE before transfer to PVDF membranes (200V for 1h). After blocking (20-min exposure to 1X Roti®-Block at room temperature), membranes were probed with relevant primary antibodies overnight at 4°C under shaking. The anti-α6 (clone MCP20), anti-α7 (clone MCP72), anti-β1 (clone MCP421), anti-PSMC2 (BML-PW8315) and anti-PSMC3 (BML-PW8310) primary antibodies were purchased from Enzo Life Sciences. Primary antibodies specific for TCF11/Nrf1 (clone D5B10), ubiquitin (clone D9D5), GAPDH (clone 14C10), PINK1 (clone D8G3), BNIP3L/NIX (clone D8G3), LC3b (#2775), eIF2α (#9722), phospho-eIF2α (ser51, #119A11) were obtained from Cell Signaling Technology. The anti-PSMD12 antibody (clone H3) was a purchase from Santa Cruz Biotechnology Inc. The anti-PA28-α (K232/1) is laboratory stock and was used in previous studies [33]. Antibodies directed 5 (ab3330), α-Tubulin (clone DM1A) and phospho-PKR (Thr446, clone E120) were purchased from Abcam. Following incubation with primary antibodies, membranes were washed three times with PBS/0.2% Tween and subsequently incubated with anti-mouse or – rabbit HRP conjugated secondary antibodies (1/5.000) for 1h at room temperature. Proteins were then visualized using an enhanced chemiluminescence detection kit (ECL) (Biorad).

### Native PAGE and proteasome in-gel peptidase activity assay

Cell pellets from resting T cell isolated from patients and related controls were lysed in ice-cold homogenization TSDG buffer (10 mM Tris pH 7.0, 10 mM NaCl, 25 mM KCl, 1.1 mM MgCl_2_, 0.1 mM EDTA, 2 mM DTT, 2 mM ATP, 1 mM NaN_3_, 20 % Glycerol) and proteins were extracted using freeze/thawing in liquid nitrogen. Protein quantification of the soluble lysates was determined using a standard bicinchoninic acid assay (BCA) from Thermo Fisher Scientific. Twenty micrograms of whole-cell lysates were run on 3-12% gradient Bis-Tris gels (Thermo Fisher Scientific) at 45V overnight at 4°C using an electrophoresis buffer consisting of 50 mM BisTris and 50 mM Tricine (pH 6,8). Following separation, peptidase activity of the proteasome was measured by incubating the gels with 0.1 mM of the suc-LLVY-AMC fluorogenic peptide (Bachem) at 37°C for 20 min in an overlay buffer (20 mM Tris, 5 mM MgCl_2_, pH 7,0). Proteasome bands were subsequently visualized by exposing the gel to UV light at 360 nm and detected at 460 nm using an Imager.

### RNA isolation, reverse-transcription and PCR analysis

Total RNA was isolated from resting T cells using the kit from Analytic Jena AG following the manufacturer’s instructions. For subsequent real-time PCR, 100-500 ng of the isolated total RNA was reverse transcribed using the M-MLV reverse transcriptase (Promega). Quantitative PCR was performed using the Premix Ex Taq™ (probe qPCR purchased from TaKaRa) and in duplicates to determine the mRNA levels of each IFN-stimulated gene (ISG) using FAM-tagged TaqMan™ Gene Expression Assays obtained from Thermo Fisher Scientific according to the manufacturer’s instructions. TaqMan™ probes used in this study for ISG quantification included *IFI27, IFI44L, IFIT1, ISG15, RSAD2, IFI44, MX1, OASL1, CXCL9* and *CXCL10*. The cycle threshold (Ct) values for target genes were converted to values of relative expression using the relative quantification (RQ) method (2^- Ct^). Target gene expression was calculated relative to Ct values for the GAPDH control housekeeping gene.

### Behavioural studies

Drosophila melanogaster (fruit flies) were raised on standard cornmeal-yeast media developed at Cold Spring Harbor Laboratory in an incubator at 23°C with 40% humidity as before [38]. Flies used for behaviour experiments included w1118 2202U (2U), Elav-Gal4. and UAS-RNAi lines targeting PMSC3 orthologues. We identified the Drosophila ortholog of PSMC3, Rpt5, using the DRSC Integrative Ortholog Prediction Tool (https://www.flyrnai.org/diopt). UAS-Rpt5 RNAi (TRiP) lines were obtained from Bloomington Drosophila Stock Center. We used the standard spatial restricted expression system Gal4- UAS to pan-neuronally express (using pan-neuronal driver Elav) the RNAi against the Drosophila ortholog Rpt5. Virgin females of Elav-Gal4 or the wild type 2U were crossed to UAS-Rpt5 males. As standard in the field, we used 2 UAS-RNAi lines to rule out the possibility of insertional or off-target effects. We included genetically appropriate controls with either UAS alone, Gal4 alone or the combination of Gal4 and UAS lines. For behavioural procedures, flies that were 0-2 days old were set up the day prior to testing and left in an incubator at 23°C with 40% humidity overnight. All testing occurred in an environmental chamber at 25°C with 70% humidity. Flies were given 1hr to acclimate to the environmental chamber prior to testing. First, Drosophila were trained to associate an odour with a foot- shock from classical olfactory conditioning as before [39]. Briefly, 100 flies are placed in a training chamber and provided with an odour (odour 1) (either octanol-OCT or methylcyclohexamide-MCH) that is presented simultaneously with a foot-shock for 60 seconds. After a brief airing, flies are then provided with a second odour (odour 2) without shock before finally being given a t-maze choice between the two odours for 2 minutes. Flies on each side of the t-maze are then counted. Then, a new set of 100 flies with the same genotype are trained to associate the other odour with a shock. A performance index is then calculated by calculating the ratio of flies avoiding the shocked odour to the total number of flies. For reversal learning, we used a similar approach to previous [40]. Reversal learning uses a similar set up and protocol with the added step of repeating the second odour 45s after it was first presented but this time paired with the shock, followed by the first odour without the shock 45s later, and finally the t-maze choice between the two odours. Statistical analysis was performed using ANOVA and then Tukey tests in JMP (SAS).

### Data representation and statistical analyses

Data are typically mean ± SEM and analyzed by pair ratio t-test between two groups. All charts and statistical analyses were generated using GraphPad Prism version 8. A p-value <0.05 was considered significant. All data are available on request from authors.

### Additional materials and methods are available in supplementary information

## Results

### Identification of *PSMC3* variants

The propositus, Subject #2, was a female newborn presenting with severe cardiac, gastrointestinal, inflammatory and immune issues. Whole-exome sequencing (WES) highlighted the *de novo* nonsynonymous c.523A>G p.(Met175Val) variant (GenBank ID: NM_002804.4), predicted to be pathogenic by bioinformatics programs and absent in any public variant databases (gnomAD, >246,000 chromosomes; NHLBI Exome Variant Server, >13,000 alleles; Bravo, 125,568 alleles). Via data sharing platform GeneMatcher [41] and direct requests in variants databases whose access was authorized to the University of Washington School of Medicine, a total of 14 distinct rare missense *PSMC3* variants could be identified *de novo* in 22 unrelated children presenting a syndrome characterized by neurodevelopmental delay (NDD) and various congenital anomalies **(Table 1)**.

**Table 1.** Main characteristics of the PSMC3 variants identified in the subjects included in the study.

| Chromosomal localization (Chr11/GRCh37) | cDNA change * | Protein change | Inheritance | Variant database ** | CADD Phred score (v1.6.) | MTR (FDR) | Metadome | MobiDetails *** | Number of subjects with the variant |
| --- | --- | --- | --- | --- | --- | --- | --- | --- | --- |
| g.47445677G>A | c.511C>T | p.(Arg171Trp) | *de novo* | rs775517283 | 27.3 | 0.573 (0.033) | I | 45364 | 1 |
| g.47445665T>C | c.523A>G | p.(Met175Val) | *de novo* | Absent | 23.4 | 0.592 (0.062) | I | 45365 | 1 |
| g.47444406G>A | c.710C>T | p.(Ala237Val) | *de novo* | Absent | 26.9 | 0.453 (0.015) | HI | 45366 | 1 |
| g.47444234T>C | c.775A>G | p.(Met259Val) | ND (suspected *de novo*) | Absent | 23.4 | 0.564 (0.061) | I | 45367 | 1 |
| g.47444233A>G | c.776T>C | p.(Met259Thr) | *de novo* | Absent | 24.7 | 0.564 (0.061) | I | 45368 | 1 |
| g.47444227A>G | c.782T>C | p.(Ile261Thr) | *de novo* | Absent | 24.6 | 0.564 (0.061) | I | 45369 | 6 |
| g.47444225C>T | c.784G>A | p.(Gly262Arg) | *de novo* | Absent | 26.8 | 0.626 (0.082) | I | 45370 | 1 |
| g.47444203C>G | c.806G>C | p.(Arg269Pro) | *de novo* | Absent | 24 | 0.622 (0.046) | I | 45371 | 1 |
| g.47444150C>G | c.859G>C | p.Glu287Gln | *de novo* | Absent | 26.7 | 0.645 (0.078) | I | 45372 | 1 |
| g.47442253G>A | c.910C>T | p.(Arg304Trp) | *de novo* | rs1363348500 | 31 | 0.392 (0.057) | I | 45373 | 4 |
| g.47442253G>C | c.910C>G | p.(Arg304Gly) | *de novo* | Absent | 28.2 | 0.392 (0.057) | I | 45374 | 1 |
| g.47442248C>A | c.915G>T | p.(Glu305Asp) | *de novo* | Absent | 23.1 | 0.357 (0.031) | I | 45375 | 1 |
| g.47442234A>G | c.929T>C | p.(Met310Thr) | *de novo* | Absent | 26.2 | 0.387 (0.05) | HI | 45376 | 1 |
| g.47440729C>T | c.1147G>A | p.(Glu383Lys) | *de novo* | Absent | 27.8 | 0.952 (0.843) | N | 45377 | 1 |

\* RefSeq transcript used for *PSMC3* is NM\_002804.4 ; \*\* gnomAD V3, dbSNP v154, ClinVar v20210828 ; \*\*\* The access to detailed predictions for variant 2288X is as follows: https://mobidetails.iurc.montp.inserm.fr/MD/api/variant/453XX/browser/MTR= Missense Tolerance Ratio; FDR= false discovery rate; Metadome: I= intolerent; HI= highly intolerent; N= neutral

As shown in **Figure 1A**, most of the *PSMC3* substitutions were localized in the “<u>A</u>TPases <u>A</u>ssociated with diverse cellular <u>A</u>ctivities” (AAA) domain and predicted to be intolerant to variations **(Figure S1)**. Two distinct regions of the AAA domain were particularly prone to substitutions. The first hotspot was centered on the recurrent variant c.910C>T p.(Arg304Trp) detected in four unrelated children and encompassed variants c.910C>G p.(Arg304Gly, c.915G>T p.(Glu305Asp) and c.929T>C p.(Met310Thr) **(Figure 1A)**. The second region enriched in rare variants [p.(Met259Thr), p.(Met259Val), p.(Ile261Thr) -seen six times-, p.(Gly262Arg) and p.(Arg269Pro)] was more N-terminally located **(Figure 1A)**. Importantly, all twelve affected residues were highly conserved across species from mammalians down to fission yeast **(Figure 1A)**.

**Figure 1:**
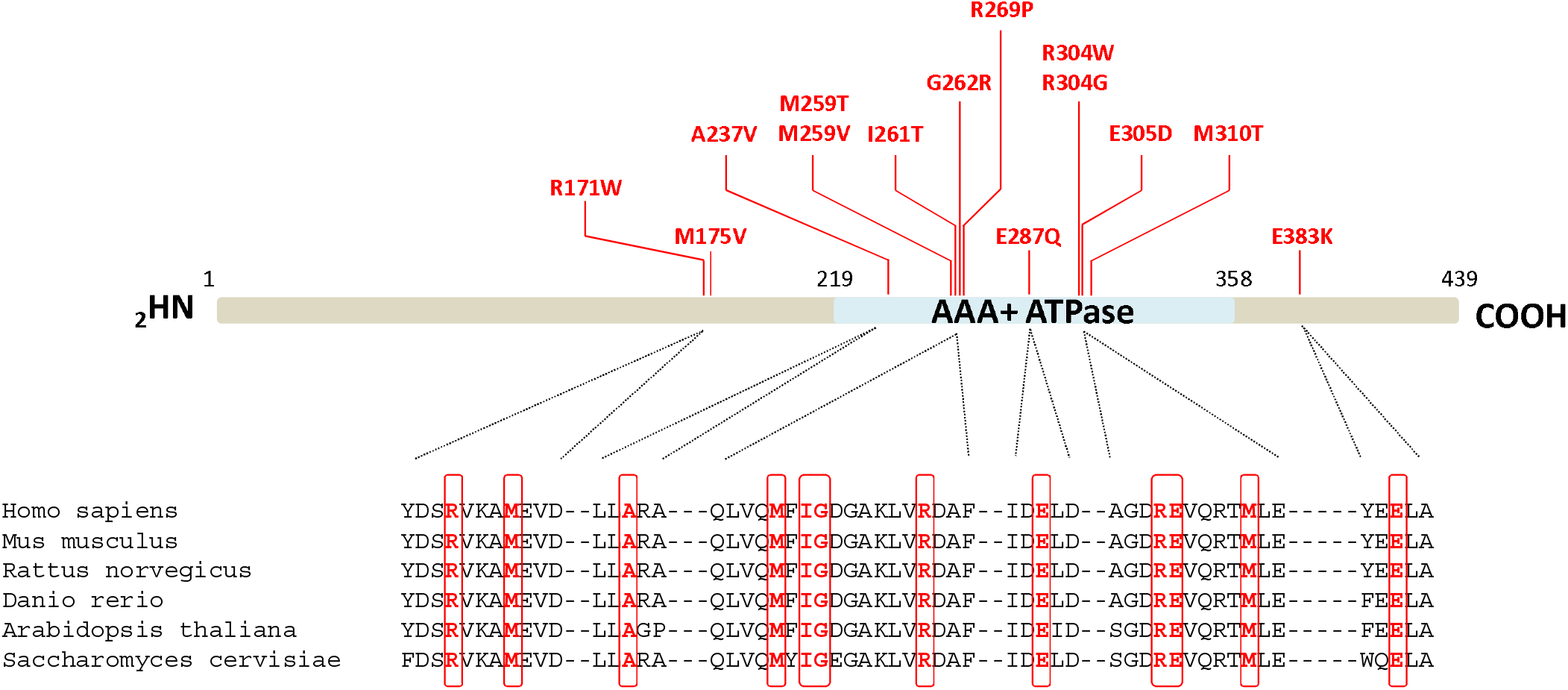
Distribution of the *de novo* heterozygous PSMC3/Rpt5 variants and morphological abnormalities in subjects affected with NDD/ID. *A.* The localization of the 13 NDD-causing missense variants in the PSMC3/Rpt5 proteins are indicated in red. The AAA-ATPase domain of the PSMC3/Rpt5 proteasome subunit of the 19S regulatory particle is depicted in blue in the schematic representation. Pink circles indicate the presence of variants hotspots. Shown is also a sequence alignment of regions immediately adjacent to the amino acids subjected to missense substitutions. Comparison of the PSMC3 primary structure across six eukaryotic organisms indicates the high conservation of the missense variant residues identified in NDD patients which are highlighted by red boxes. *B.* Facial photos of affected subjects were withdrawn from the present version of the manuscript but can be accessed by querying GestaltMatcher database (https://db.gestaltmatcher.org/) using the term “PSMC3” in the gallery section.

One major phenotype hallmark of all individuals with *PSMC3* variants is the predominance of neurodevelopmental or neuropsychiatric symptoms **(Table 2)**. In more details, apart from Subject #2, all affected children had developmental delay (21/21; 100%) characterized by speech delay (18/18; 100%) and/or intellectual disability (15/17; 88%), and motor delay (14/18; 78%). Brain magnetic resonance imaging highlighted frequent anomalies (11/14; 79%), whereas abnormal behavior (9/17; 53%) and seizures (5/20; 25%) were inconstant. 9/18 (50%) individuals experienced growth failure, with feeding difficulties (8/17; 47%). Malformations were frequently observed notably in skeleton (10/14; 71%; scoliosis, acetabular dysplasia, brachymetatarsy), heart (10/17; 59%; ventricular or septal defects, patent ductus arteriosus, pulmonary hypertension and atresia), kidney (4/14; 29%; horseshoe shape, pelvicalyceal dilatation, nephrocalcinosis, and multi-cystic dysplastic kidney), and head (microcephaly in 6/16 (38%); relative to severe macrocephaly in 2/16 (13%)). Tumors were noted in 2/18 (11%) individuals (craniopharyngioma and neuroblastoma). Hearing loss was detected in 8/18 individuals (44%) and labeled as sensorineural in two and conductive in one of them, respectively. Most children (17/19; 89%) displayed dysmorphic facial features, including notably tall or broad forehead (7/19; 37%), thin upper lip with down-turned corners of mouth (6/19; 32%), abnormal palate (5/19; 265/19; 26%), epicanthal folds (5/19; 26%), and orofacial clefts (2/19; 10%). Computational analysis of facial morphology by GestaltMatcher [42] revealed that facial dysmorphism among the *PSMC3* subjects was rather heterogeneous with similarities only observed between patients carrying identical variants **(Figure S2).**

**Table 2.** Phenotype of subjects with de novo PSMC3 variants (1/3)

| Subject | #1 <sup>a</sup> | #2 | #3 <sup>b</sup> | #4 | #5 | #6 | #7 |
| --- | --- | --- | --- | --- | --- | --- | --- |
| Variant in *PSMC3* | c.511C>T | c.523A>G | c.710C>T | c.775A>G | c.776T>C | c.782T>C | c.782T>C |
|  | p.(Arg171Trp) | p.(Met175Val) | p.Ala237Val | p.(Met259Val) | p.(Met259Thr) | p.(Ile261Thr) | p.(Ile261Thr) |
| Gender | Male | Female | Male | Female | Male | Female | Female |
| Developmental delay | + | N/A | + | + | + | + | + |
| Speech delay | + | N/A | N/A | + | + | + | N/A |
| Facial dysmorphism | N/A | N/A | + | + | + | + | + |
| Intellectual disability | N/A | N/A | + | + | + | + | N/A |
| Brain MRI anomalies | N/A | + | N/A | N/A | - | + | + |
| Motor delay | N/A | N/A | N/A | - | + | + | + |
| Skeletal malformations | N/A | N/A | N/A | - | + | - | + |
| Gastrointestinal problems | N/A | + | N/A | - | + | - | - |
| Cardiac malformations | N/A | + | + | - | + | - | N/A |
| Hypotonia/abnormal tone | N/A | N/A | N/A | + | + | - | - |
| Abnormal behavior | + | N/A | N/A | - | + | + | - |
| Growth failure | N/A | + | N/A | - | - | - | - |
| Feeding difficulties | N/A | + | N/A | - | + | - | - |
| Hearing loss | N/A | N/A | N/A | + | + | - | - |
| Microcephaly | N/A | - | N/A | - | - | - | N/A |
| Other malformations | N/A | N/A | N/A | - | + | N/A | - |
| Genital abnormalities | N/A | N/A | N/A | - | + | - | - |
| Renal malformations | N/A | N/A | - | - | + | - | - |
| Seizures | N/A | - | + | - | + | - | - |
| Underweight | N/A | + | N/A | - | - | - | - |
| Short stature | N/A | - | N/A | - | N/A | - | - |

\* Nomenclature HGVS V2.0 according to mRNA reference sequence NM\_002804.4. Nucleotide numbering uses +1 as the A of the ATG translation initiation codon in the reference sequence, with the initiation codon as codon 1. N/A: parameter not analyzed; <sup>a</sup>information retrieved from 'Simons Foundation Powering Autism Research' (SPARK) cohort; <sup>b</sup>information retrieved from 'Deciphering Developmental Disorders' (DDD) cohort.

**Table 2.** Phenotype of subjects with *de novo PSMC3* variants (2/3)

| Subject | #8 | #9 | #10<sup>b</sup> | #11 | #12 | #13 | #14<sup>b</sup> | #15 |
| --- | --- | --- | --- | --- | --- | --- | --- | --- |
| Variant in *PSMC3* | c.782T>C | c.782T>C | c.782T>C | c.782T>C | c.784G>A | c.806G>C | c.859G>C | c.910C>T |
|  | p.(Ile261Thr) | p.(Ile261Thr) | p.(Ile261Thr) | p.(Ile261Thr) | p.(Gly262Arg) | p.(Arg269Pro) | p.Glu287Gln | p.(Arg304Trp) |
| Gender | Male | Male | Male | Male | Male | Female | Male | Male |
| Developmental delay | + | + | + | + | + | + | + | + |
| Speech delay | + | + | + | + | + | + | N/A | + |
| Facial dysmorphism | + | + | + | + | - | N/A | + | + |
| Intellectual disability | + | + | + | + | - | + | N/A | + |
| Brain MRI anomalies | - | N/A | + | N/A | - | + | + | + |
| Motor delay | + | - | + | - | - | + | N/A | + |
| Skeletal malformations | + | N/A | + | - | N/A | N/A | N/A | + |
| Gastrointestinal problems | + | + | + | + | - | - | N/A | + |
| Cardiac malformations | - | N/A | - | - | + | N/A | + | + |
| Hypotonia/abnormal tone | + | + | + | - | - | + | N/A | + |
| Abnormal behavior | + | - | - | + | - | N/A | N/A | - |
| Growth failure | - | - | - | - | + | N/A | N/A | + |
| Feeding difficulties | - | - | - | + | - | N/A | N/A | + |
| Hearing loss | - | - | - | + | + | + | - | + |
| Microcephaly | - | - | N/A | - | - | N/A | N/A | + |
| Other malformations | - | - | + | - | N/A | N/A | + | - |
| Genital abnormalities | - | + | + | - | N/A | N/A | - | + |
| Renal malformations | N/A | N/A | N/A | - | N/A | N/A | - | + |
| Seizures | - | - | + | + | - | - | - | + |
| Underweight | - | - | - | - | + | N/A | N/A | - |
| Short stature | - | - | - | - | - | - | N/A | + |

**Table 2.** Phenotype of subjects with de novo PSMC3 variants (3/3)

| Subject | #16 | #17 | #18<sup>b</sup> | #19<sup>b</sup> | #20 | #21 | #22 | Total<br>(22 patients) |
| --- | --- | --- | --- | --- | --- | --- | --- | --- |
| Variant in *PSMC3* | c.910C>T | c.910C>T | c.910C>T | c.910C>G | c.915G>T | c.929T>C | c.1147G>A |  |
|  | p.(Arg304Trp) | p.(Arg304Trp) | p.(Arg304Trp) | p.(Arg304Gly) | p.(Glu305Asp) | p.(Met310Thr) | p.(Glu383Lys) |  |
| Gender | Male | Female | Female | Female | Female | Male | Male | 13M/9F |
| **Developmental delay** | + | + | + | + | + | + | + | **21/21 (100%)** |
| **Speech delay** | + | + | + | + | + | + | + | **18/18 (100%)** |
| **Facial dysmorphism** | + | + | + | + | + | - | + | **17/19 (89%)** |
| **Intellectual disability** | + | + | + | + | N/A | + | - | **15/17 (88%)** |
| **Brain MRI anomalies** | + | + | N/A | + | N/A | N/A | + | **11/14 (79%)** |
| **Motor delay** | + | + | + | + | + | + | + | **14/18 (78%)** |
| **Skeletal malformations** | + | + | N/A | + | + | - | + | **10/14 (71%)** |
| Gastrointestinal problems | + | N/A | N/A | - | - | + | + | 10/17 (59%) |
| Cardiac malformations | + | + | + | + | N/A | - | - | 10/17 (59%) |
| Hypotonia/abnormal tone | + | + | N/A | + | - | - | - | 10/17 (59%) |
| Abnormal behavior | + | - | N/A | + | + | + | - | 9/17 (53%) |
| Growth failure | + | + | + | - | + | + | + | 9/18 (50%) |
| Feeding difficulties | + | + | N/A | + | - | - | + | 8/17 (47%) |
| Hearing loss | + | + | N/A | - | - | - | - | 8/18 (44%) |
| Microcephaly | + | + | + | - | + | + | - | 6/16 (38%) |
| Other malformations | + | N/A | N/A | - | - | - | + | 5/14 (36%) |
| Genital abnormalities | - | + | N/A | - | - | - | - | 5/16 (31%) |
| Renal malformations | + | + | N/A | - | - | - | - | 4/14 (29%) |
| Seizures | - | - | N/A | - | - | - | - | 5/20 (25%) |
| Underweight | - | - | N/A | - | + | - | + | 4/17 (24%) |
| Short stature | - | - | N/A | - | + | - | + | 3/17 (18%) |

### *PSMC3*-silenced *Drosophila* adult flies fail to reverse stimulus contingencies

Given the neuronal nature of the phenotype of *PSMC3* subjects, we next sought to address the potential involvement of *PSMC3* in cognitive function by evaluating the learning performance of Drosophila fruit flies with a knockdown of *PSMC3* (i.e. Rpt5) expression. To this end, we used a standard conditioning of odour-avoidance response in which animals were exposed to two different odours (OCT or MCH), only one of which resulting in the simultaneous application of a foot-shock (OCT+, MCH-), as previously described [39] **(Figure 2A)**. As shown in **Figure 2C**, pan-neuronal expression of Rpt5 RNAi resulted in no significant differences in learning performance for Rpt5^32422^ (P=0.6435, N=4). Similarly, RNAi expression in Rpt5^53886^ resulted in normal learning performance (P=0.5282, N=6) as well. We next determined the reversal learning performance of RPT5 silenced flies by training with an initial odour shock pairing (e.g. OCT+, MCH-) immediately followed by training with a reversed odour shock pairing (e.g. OCT-, MCH+) **(Figure 2B)**. Strikingly, reversal learning performance was significantly defective with pan-neuronal Rpt5 RNAi expression for Rpt5^32422^ (P<0.0001, N=4); the three control groups did not significantly differ from each other. RNAi expression also led to defective reversal learning performance for Rpt5^53886^ (P=0.0022, N=6); the three control groups once again did not significantly differ from each other **(Figure 2B)**. These data suggest that *PSMC3* appears as a prerequisite for the changes in learned associations.

**Figure 2:**
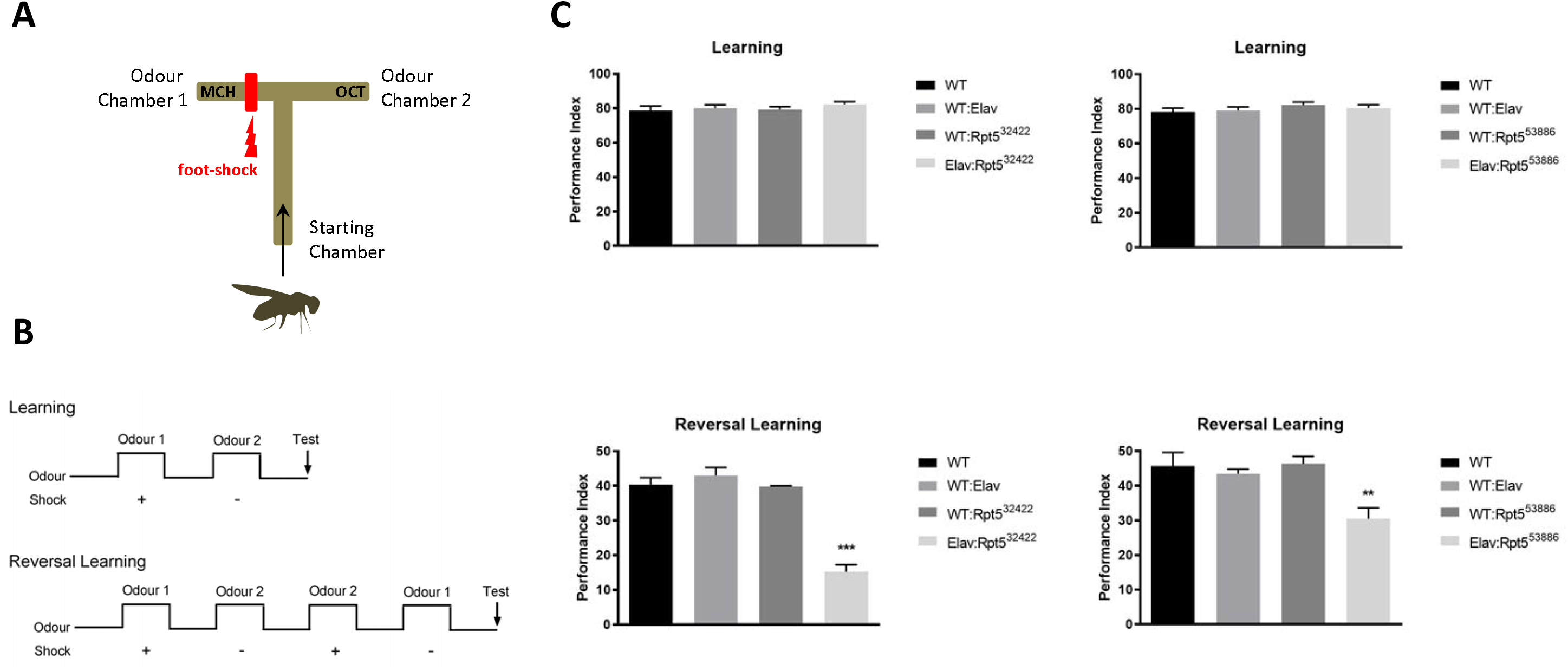
Pan-neuronal disruption of PSMC3/Rpt5 results in normal learning performance but defective reversal performance. ***A***. A T-maze, depicting standard conditioning of odour-avoidance response in which Drosophila flies are trained to avoid one particular odour chamber which is associated with a foot-shock (in this example, odour chamber 1). ***B***. Schematic illustration of the experimental timelines of the learning and reversal learning protocols used in these experiments. Reversal learning is assessed by reversing odour shock pairing (MCH-, OCT+), as indicated. ***C***. Upper left: no significant difference was detected in learning with pan-neuronal expression of Rpt5 RNAi for Rpt5^32422^ (P=0.6435, N=4). Upper right: similarly, pan-neuronal expression of Rpt5 RNAi for Rpt5^53886^ resulted in no significant difference in learning (P=0.5282, N=6). Lower left: reversal learning was significantly defective with pan-neuronal Rpt5 RNAi expression for Rpt5^32422^ (P<0.0001, N=4). Lower right: pan-neuronal Rpt5 RNAi expression for Rpt5^53886^ also resulted in significant learning defects (P=0.0022, N=6).

### Ectopic expression of PSMC3/Rpt5 or its variants differentially impact neuronal development

In view of the negative impact of *PSMC3* gene silencing on reversal learning, we next asked whether *PSMC3* was involved in the regulation of hippocampal neuron dendritic development. To address this point, we ectopically expressed wild-type *PSMC3* in murine primary hippocampal neurons prior to neurite length quantification, as previously described [43]. As shown in **Figure S3**, expression of wild-type *PSMC3* at an early developmental time point in vitro (Day In Vitro (DIV) 3) resulted in significantly reduced neurite length of the neurons when fixed 5 days later, at DIV8. These results indicate that increased levels of *PSMC3* is damaging for neuronal development. We next sought to determine whether the different *PSMC3* variants identified in NDD/ID patients behave differently compared to PSMC3 WT in neurons when ectopically expressed. Interestingly, expression of the p.(Arg304Trp), p.(Glu305Asp) and p.(Glu383Lys) variants resulted in similar neuronal morphological changes as seen with wild-type *PSMC3* **(Figure S3)**. By contrast, expression of the p.(Met175Val) variant did not affect neuronal morphology when compared to the empty vector control, and showed significant improvement when compared to wild-type *PSMC3*. Taken together these results suggest that *PSMC3* participates in the regulation of neurite development and that any alteration of this gene might affect this process positively or negatively.

### *PSMC3* variants differentially affect the Rpt5 steady-state protein level

Because missense variants may cause haploinsufficiency by affecting mRNA and/or protein turnover, we next sought to determine the impact of the identified *PSMC3* variants on the Rpt5 steady-state expression level. To this end, nine *PSMC3* point variants were introduced in the SHSY5Y neuroblastoma cell line and expressed with a fused N-terminal HA tandem repeat prior to western-blot analysis. As shown in **Figure 3A**, four variants including p.(Arg171Trp), p.(Ala237Val), p.(Met259Val) and p.(Ile261Thr) had substantially lower steady-state protein levels than their wild-type counterpart. However, all *PSMC3* variants generated equivalent amounts of PSMC3 transcripts in SHSY5Y cells following a 24-h transfection **(Figure 3B)**, indicating that reduced expression at the protein level was due to increased protein turnover and/or decreased translation efficiency. Altogether, these data show that the *PSMC3* missense variants identified in NDD/ID subjects do not necessarily behave similarly.

**Figure 3:**
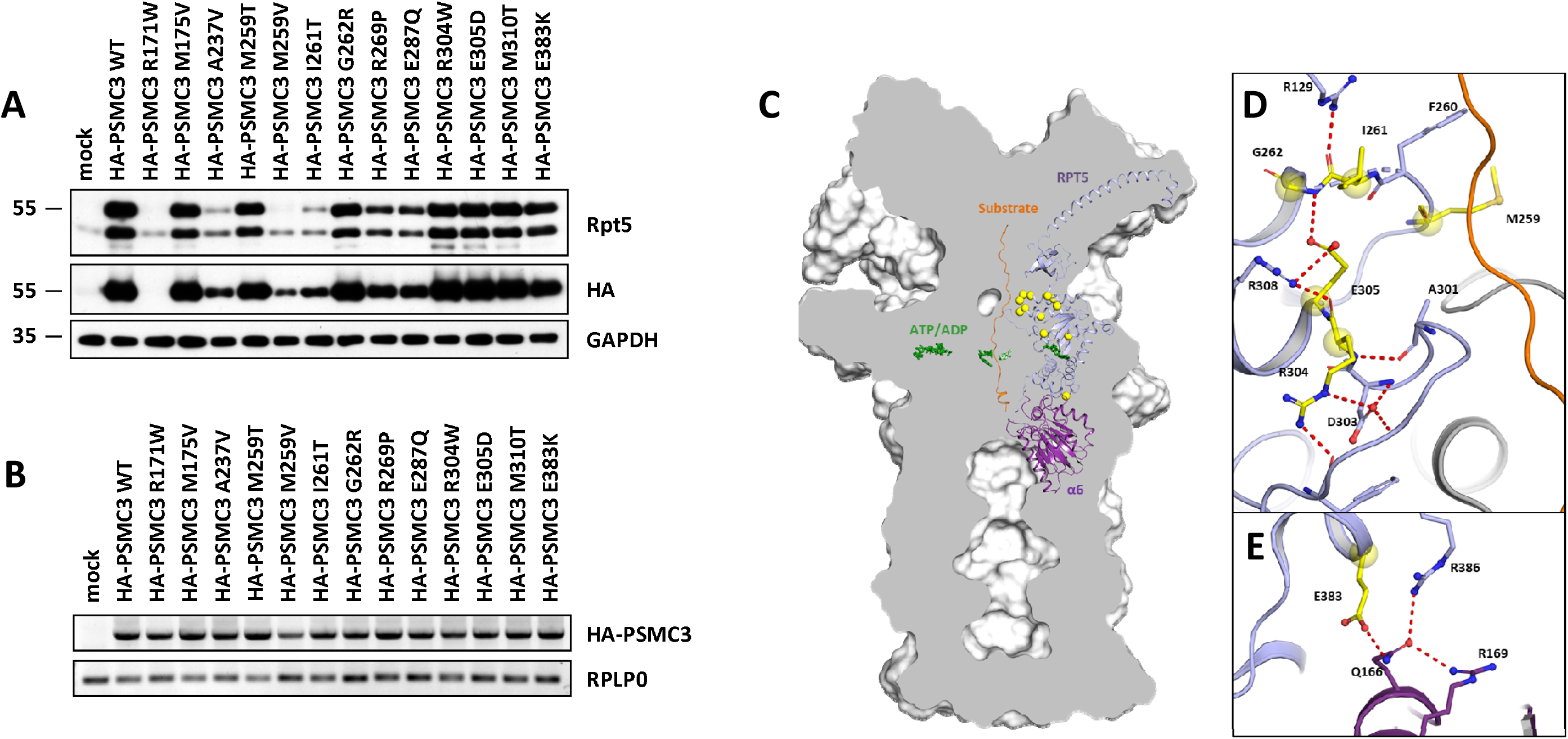
PSMC3/Rpt5 variants do not behave similarly at the molecular level. ***A*.** SHSY5Y cells were transfected with HA-tagged *PSMC3* mutants for 24 h prior to protein extraction and western-blotting using antibodies specific for PSMC3 and HA, as indicated. Both non-transfected and mock-transfected cells served as negative controls. Equal protein loading was ensured by probing the membranes with an anti-α-tubulin monoclonal antibody. Shown is one representative experiment out of two. ***B*.** SHSY5Y cells transfected with the various HA-PSMC3 variants were subjected to total RNA extraction followed by semi-quantitative RT-PCR using primer located in PSMC3 and the polyadenylation signal of the pcDNA3.1/*myc*-HIS expression vector (BGH). Equal loading between the samples was ensured by amplifying the RPL0 gene, as indicated. ***C*.** A sliced surface view of the 26S proteasome (grey) was superimposed with a cartoon representation of the subunits PSMC3/RPT5 (blue) and PSMB1/α6 (purple) as well as the substrate (orange). The ATP/ADP molecules of the AAA-ATPase ring are shown as green sticks, while the investigated missense variants as bright yellow spheres. ***D*.** Detailed representation of the missense variants in the loop region of the N-terminal α/β domain. Residues affected byvariants are involved in a polar interaction network close to the substrate tunnel. ***E*.** Close up view on the RPT5-α6 interface affected by the p.(Glu383Lys) variant. Residues affected by variants are shown as bright yellow balls and sticks with atoms coloured by polarity (oxygen in red, nitrogen in blue and sulphur in dark yellow). Figures were created with PyMOL v. 2.0 (pymol.org) using the human 26S proteasome structure (PDB-entry code: 6MSK [44]).

### *PSMC3* substitutions are predicted to affect inter and intra-molecular interactions between proteasome subunits

We next attempted to predict the structural consequences of the each of the fourteen *PSMC3* substitutions by assessing the localization of the mutated residues in the human 26S proteasome structure generated by Dong et al. (PDB-entry code: 6MSK) [44]. As shown in **Figure 3C**, most of the affected amino acids emerge within the N-terminal α/β domain of PSMC3/Rpt5 with five residues (i.e. Gly262, Ile261, Met259, Arg304, Glu305) residing in two loops adjacent to the substrate tunnel pointing towards the center of the AAA-ATPase ring **(Figure 3D)**. Specifically, on one loop, Gly262 is fixed by a main chain hydrogen bond to Glu305, thereby promoting flexibility of the preceding loop containing Met259. Besides, Glu305 itself is held through a salt bridge by Arg308 with its preceding residue Arg304 involved in a polar network stabilizing the neighbouring loop **(Figure S4)**. Because these six residues stabilize or are part of the tertiary structure of the loops, any alteration of these amino acids is predicted to affect substrate trafficking, as well as interactions with other AAA-ATPase subunits. As shown in **Figure S4**, the Ala237Val variant is more difficult to classify and does not reveal itself on structural level at first sight. However, one cannot exclude that the slight increase in residue size at position 237 might lead to structural changes. Indeed, our overexpression assays in SHSY5Y cells suggest that such substitution does affect sidechain packing and protein stability **(Figure 3A)**. The p.(Glu383Lys) missense variant is the only substitution lying within the C-terminal α-helical domain of PSMC3/Rpt5 adjacent to the 19S-20S interface. Interestingly, Glu383 holds Gln166 from the PSMA1/α6 subunit for polar interactions with both of the Arg169 and Arg386 residues **(Figure 3E)**. Variant of the negatively charged Glu383 to a positively charged Lys383 is therefore predicted to disrupt such hydrogen bond network and affect the association of the 19S complex with the 20S core particle. Interestingly, Glu287 is located in close proximity of the ATP binding site within PSMC3/Rpt5 **(Figure S4)** and its substitution into Gln287 presumably generates additional polar bonds with Asn333 likely to affect ATP binding and/or hydrolysis. As illustrated in **Figure S5**, Arg171 is positioned at the PSMC3/Rpt5-PSMC2/Rpt1 interface and is part of a polar network involving the neighboring Asp169 and Asn258 residues. As such, the substitution of positively charged arginine to hydrophobic tryptophan at this position is predicted to disrupt these interactions, and a fortiori to affect the contact between the two subunits **(Figure S5)**. As for the Met175Val residue, its change to Val175 results in the loss of a polarized thiol group and hydrophilic environment is likely to destabilize the tertiary structure of this protein region **(Figure S5)**. Taken together, these data suggest that the complex 26S proteasome structure could be strongly affected by the identified PSMC3/Rpt5 missense variants through distinct mechanisms including substrate translocation or 19S- ATPase-interaction with the 20S particle.

### *PSMC3* variants differentially impact proteasome assembly in NDD/ID subjects

To further address the pathogenicity of the *PSMC3* variants, T cells from Subjects #12, #17 and #20, were next analyzed by SDS-PAGE/western-blotting for their proteasome contents. As shown in **Figure 4A**, the expression levels of the proteasome α-subunits (i.e. α7) and PA28-α did not significantly vary between controls and index cases. Likewise, the abundance of the 19S subunits (i.e. Rpt5 and Rpn5) in mutant T cells was quite comparable to that detected in their control counterparts. Most importantly, none of *PSMC3* variant T cells showed reduced expression of the Rpt5 full-length protein, suggesting that proteasome dysfunction in these affected individuals was not due to haploinsufficiency.

**Figure 4:**
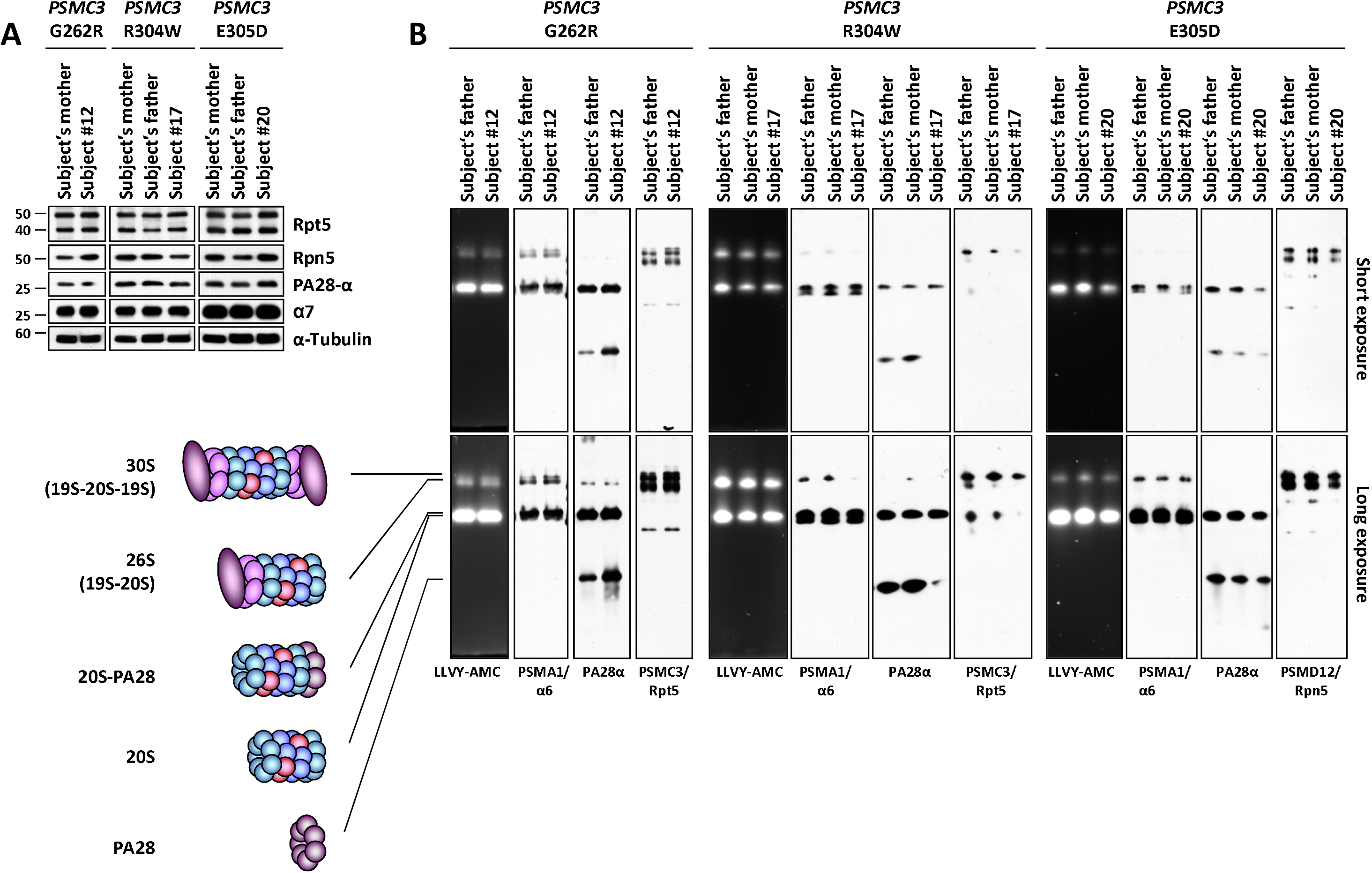
PSMC3/Rpt5 variants cause proteasome assembly defects in individuals with NDD/ID. ***A*.** Five to twenty micrograms of RIPA lysates from T cells isolated from Subjects #12, #17 and #20 as well as related controls (index case’s father and/or mother) were separated by SDS-PAGE followed by western-blotting using antibodies directed against PSMC3/Rpt5, PSMD12/Rpn5, PA28-α and Alpha7, as indicated. Equal protein loading was ensured by probing the membrane with an anti-α-tubulin antibody. ***B*.** Twenty micrograms of native whole-cell lysates derived from T cells isolated from NDD Subjects #12, #17 and #20 as well as related controls (index case’s father and/or mother) were separated by 3-12% native-PAGE prior to suc-LLVY-AMC in-gel activity assay to visualize the positions of the 20S and 26S proteasome complexes. Gels were subsequently transferred to blots and probed with antibodies specific for Alpha6, PSMC3/Rpt5, PSMD12/Rpn5 and PA28-α indicated.

T cells from affected individuals and relative controls (i.e. father and/or mother) were next analyzed for proteasome complex formation and activity by in-gel fluorescence or western blotting on native PAGE. As shown in **Figure 4B**, the chymotrypsin-like activity of the 26S and 20S proteasome complexes was reduced in Subjects #17 and #20, respectively, while it was mostly unchanged in Subject #12. Subsequent western-blot analysis revealed that the decreased 20S activity observed in Subject #20 was due to a decreased pool of her 20S-PA28 complexes, as determined by reduced band intensity for the α6 and PA28 proteins. In a similar fashion, a diminished expression of the α6, PSMC3/Rpt5 and PSMD12/Rpn5 subunits was observed in the 26S proteasomes of Subject #17 **(Figure 4B)**, indicating that the decline in 26S activity detected in this patient was likely to be attributed to decreased amounts of 26S complexes. However, the 20S and 26S proteasome pools of Subject #12 did not substantially vary when compared to those of his related control **(Figure 4B).** Altogether, these data indicate that both of the p.(Arg304Trp) and p.(Glu305Asp) PSMC3/Rpt5 variants critically affect 20S and/or 26S proteasome assembly in individuals with NDD/ID, while the p.(Gly262Arg) variant has little impact in this process.

### Quantitative proteomics identifies specific biomarkers of *PSMC3* loss-of-function in NDD/ID subjects

To better understand the cellular consequences of *PSMC3* loss-of-function, we next performed a mass spectrometry (MS)-based comparative analysis of the proteome of Subjects #16 and #20 (p.Arg304Trp and p.Glu305Asp, respectively) to that of their wild-type counterparts. As shown in **Figure 5**, our data identified a protein biomarker signature consisting of seventeen ribosomal proteins of the small 40S (i.e. RPS) or large 60S (i.e. RPL) ribosomal subunits which were specifically upregulated in both investigated patients. This suggests that mRNA translation is a major affected pathway upon *PSMC3* loss-of-function, a notion which is further supported by the view that components of the mRNA processing machinery such as CELF1, LSM1, SSU72 and ITPA are also differentially expressed across patients and controls **(Figure 5)**. Other notable proteins whose abundances vary in *PSMC3* subjects include components of the immune system such as MX1 –a typical interferon (IFN)-stimulated gene product– and the α-chain of the IL3 receptor (IL3RA). Interestingly, these proteins are regulated in opposite directions with both patients exhibiting higher amounts of MX1 but reduced levels of IL3RA **(Figure 5)**. Our analysis further revealed that *PSMC3* loss-of-function was also associated with increased protein levels of the H1.5 and H1.2 linker histone H1 variants, a finding that may point to a distinct chromatin regulation in these patients. Collectively, these data suggest that patients with *PSMC3* variants exhibit alterations in basic cellular processes including mRNA translation, immune signaling and chromatin remodeling.

**Figure 5:**
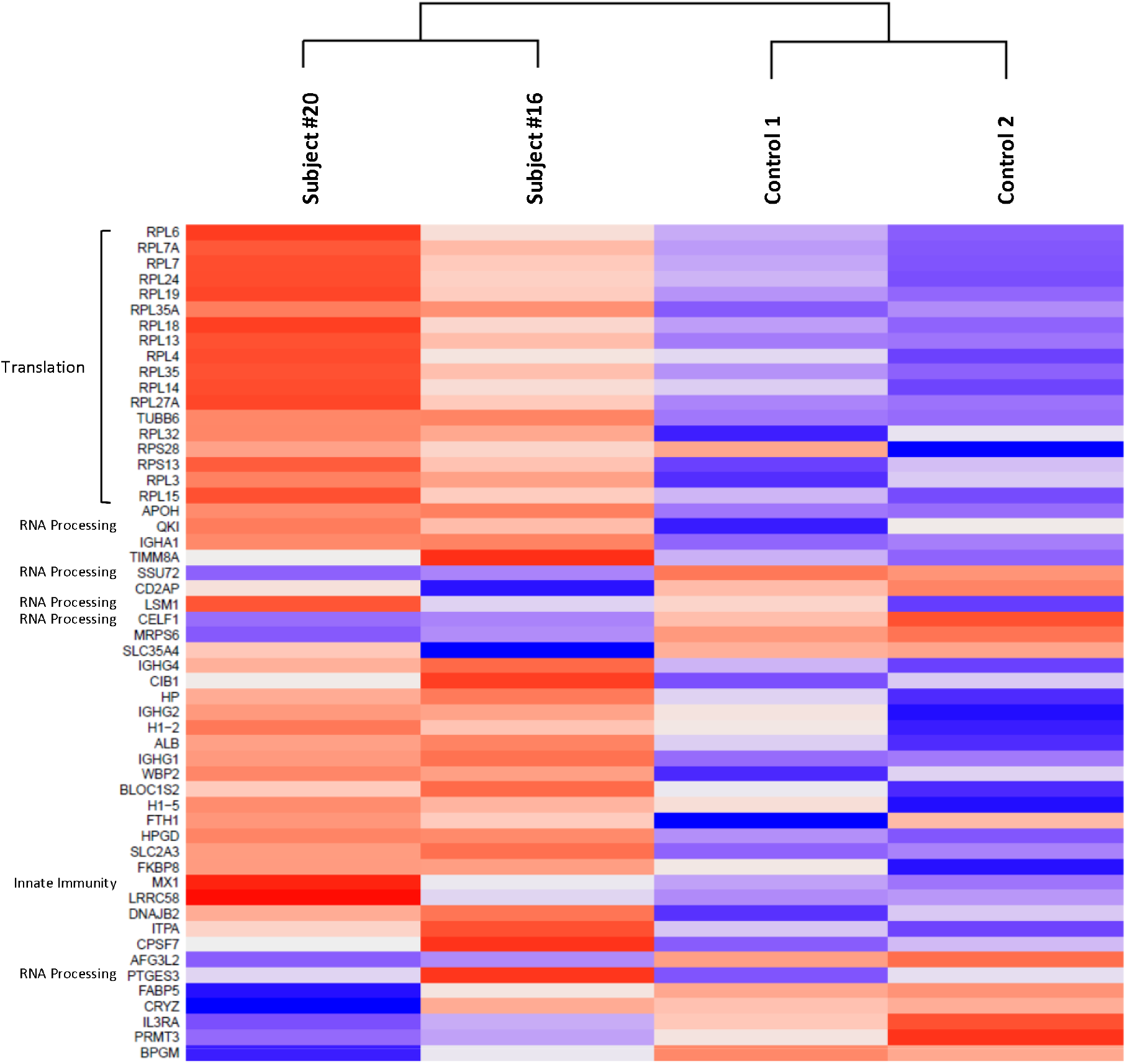
Proteomic signatures of NDD/ID subjects carrying PSMC3/Rpt5 variants. Heatmap cluster analysis showing the similarities in the protein expression profiles of the *PSMC3* Subjects #16 and #20 (carrying the p.Arg304Trp and Glu305Asp *PSMC3* variants, respectively) compared to their related controls (father and/or mother of the proband), as indicated. Only the differentially expressed proteins with an absolute value of log2 fold-change greater than 2 were selected for the clustering analysis.

### *PSMC3* variants cause proteotoxic stress in patient cells

Control and subject T cells were next analyzed for their content in ubiquitin-protein conjugates by western-blotting. As shown in **Figure 6A**, all four investigated patients exhibited typical features of unbalanced protein homeostasis, as evidenced by increased accumulation of ubiquitin-modified species when compared to their respective related controls.

**Figure 6:**
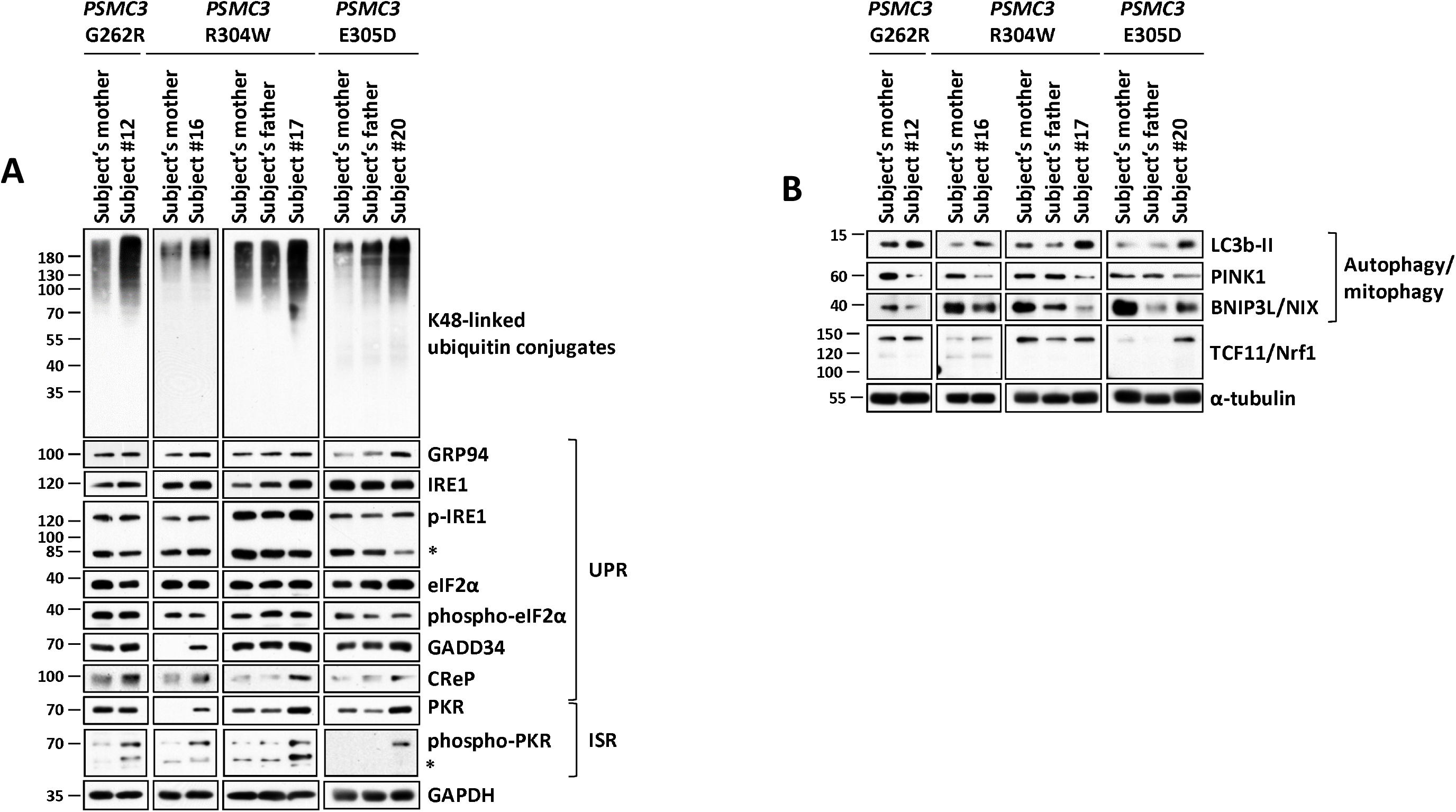
NDD/ID subjects carrying PSMC3/Rpt5 variants exhibit severe signs of protein homeostasis perturbations and alterations of the UPR, ISR and autophagy/mitophagy pathways. ***A*.** Five to twenty micrograms of RIPA lysates from T cells isolated from subjects #12, #16, #17 and #20 as well as related controls (*PSMC3* index case’s father and/or mother) were separated by SDS-PAGE followed by western-blotting using antibodies directed against K48-linked ubiquitin-modified proteins, GRP94, IRE1, phospho-IRE1, eIF2α, phospho-eIF2α, GADD34, CReP, PKR, phospho-PKR and GAPDH (loading control), as indicated. ***B*.** RIPA-cell lysates from Subjects #12, #16, #17 and #20 as well as their related controls (*PSMC3* index case’s father and/or mother) were subjected to SDS-PAGE/western-blotting using antibodies specific for LC3b, PINK1, BNIP3L and α tubulin (loading control), as indicated.

Proteotoxic stress is known to induce the unfolded and integrated stress response (i.e. UPR and ISR, respectively) [26]. To address this point, we monitored in both control and variant T cells from Subjects #12, #16, #17 and #20 the expression level of the GRP94 chaperone protein whose upregulation is understood to be a major hallmark of the UPR [45]. Indeed, as shown in **Figure 6A**, all four investigated *PSMC3* subjects exhibited increased steady-state expression levels of GRP94 when compared to their respective related controls, indicating that their T cells suffer from ER stress. However, the activation of the UPR was only partial, as the phosphorylation/activation status of two other UPR markers, namely IRE1 and eIF2α was not changed between controls and *PSMC3* subjects **(Figure 6A)**. The failure to detect increased phosphorylated eIF2α is intriguing considering the fact that one its major upstream kinases, protein kinase R (PKR), was consistently activated in all subjects **(Figure 6A)**. It may however be easily explained by the observation that both eIF2α phosphatases GADD34 and CReP were consistently up-regulated across all four patients.

Interestingly, T cells with *PSMC3* variants were also endowed with increased steady-state expression levels of LC3-II **(Figure 6B)**, suggesting that the inability of these cells to eliminate ubiquitin-protein aggregates cells via their 26S proteasomes triggers a compensatory mechanism mediated by activation of the autophagy system. Consistently, the mitochondrial proteins PINK1 and Bnip3L/NIX were found to be decreased in all four NDD/ID subjects with *PSMC3* variants **(Figure 6B)**, supporting the notion that selective autophagic processes including mitophagy were persistently activated upon *PSMC3* disruption. Because proteasome impairment typically results in the release of the TCF11/Nrf1 transcription factor from the ER membrane [46, 47], we next sought to determine whether TCF11/Nrf1 was processed in NDD/ID affected individuals. However, no difference in the TCF11/Nrf1 processing pattern could be detected between controls and *PSMC3* subjects **(Figure 6B)**, suggesting that *PSMC3* variants associated with NDD/ID do not initiate the TCF11/Nrf1 signaling pathway.

### T cells isolated from patients harboring *PSMC3* variants exhibit a typical type I IFN signature

Because proteasome loss–of–functions caused by variants in genes of 20S core particle subunits result in the generation of a typical type I IFN response [32], we next sought to determine whether alterations in the *PSMC3* gene would induce interferonopathies as well. To this end, we undertook a comparative examination of the mRNA levels of 750 predefined immunological relevant genes in T cells from *PSMC3* subjects and relative controls (father and/or mother) using the NanoString® nCounter platform. Interestingly, a total of 30 differentially expressed genes could be identified including 11 interferon-stimulated genes (ISG) which were specifically upregulated in all NDD/ID patients **(Figure 7A)**, suggesting that *PSMC3* loss-of-function is associated with a type I IFN response. Besides, our transcriptomic analysis of control and subject T cells further revealed that PSMC3 disruption resulted in the upregulation of genes of the notch signaling pathway such as NOTCH2 and JAG2 involved in neurodevelopment [48] **(Figure 7A)**.

**Figure 7:**
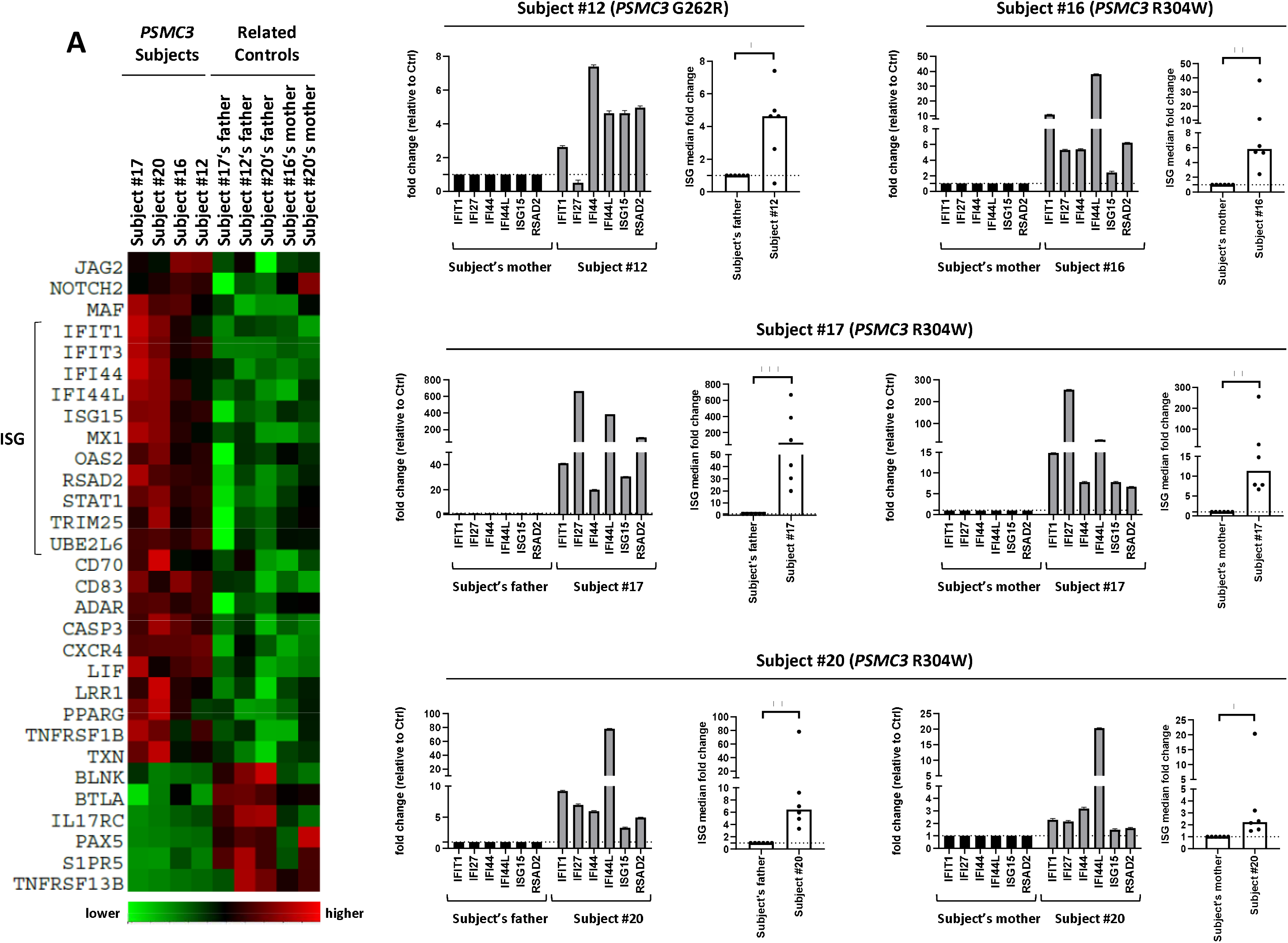
NDD/ID subjects with PSMC3/Rpt5 missense variants exhibit a typical type I interferon (IFN) signature. ***A*.** Heat map clustering of gene expression in T cells isolated from *PSMC3* subjects and their relative controls (father and/or mother of the proband). Each column represents one individual patient/related control and each row represents one gene. Clustering of genes and samples was carried out by centred Pearson correlation. Colour indicates normalized counts of each gene, with green representing higher expression and red relatively lower expression. ***B*.** Gene expression of six typical IFN-stimulated genes (*IFIT1*, *IFI27*, *IFI44*, *IFI44L*, *ISG15* and *RSAD2*) was assayed by RT-qPCR on T cells derived from the NDD/ID Subjects #12 (A), #16 (B), #17 (C) and #20 (D) as well as their respective controls (*PSMC3* index case’s father and/or mother). Expression levels were normalized to GAPDH and relative quantifications (RQ) are presented as fold change over controls. Shown is also the median fold expression of the six ISG over relative controls. Statistical significance was assessed by ratio paired t test where *indicates p<0.05, ** indicates p<0.01 and *** indicates p<0.001.

To validate the type I IFN gene signature unraveled by our omics profiling of the *PSMC3* subjects, we next evaluated the transcription rate of six ISG (*IFI27*, *IFI44L*, *IFIT1*, *ISG15*, *RSAD2* and *IFI44*) by quantitative PCR in T cells isolated from these four families with *PSMC3* missense variants. Strikingly, as shown in **Figure 7B**, all four affected children (Subjects #12, #16, #17 and #20) exhibited much higher ISG expression levels than their parents (father and/or mother) used as controls. Calculation of the median fold change of the six ISG revealed that a minimum and significant increase of 2.23-fold was detected in all *PSMC3* index cases when compared to their respective controls. The strongest type I IFN signature was observed in Subject #17 whose ISG were upregulated by approximately 75- and 12-fold when compared to her father and mother, respectively. Subjects #12, #16 and #20 exhibited a milder type I IFN induction characterized by a 2-to 6-fold greater amount of ISG transcripts than their respective controls. Among the six ISG tested, IFIT1, and IFI44L were the genes which underwent the most pronounced upregulation in all four NDD/ID affected individuals.

To strengthen our view that NDD/ID affected individuals with *PSMC3* missense variants develop an interferonopathy, we next calculated and compared the IFN scores of both *PSMC3* subjects and their related controls to those of T cells isolated from six healthy donors. As shown in **Figure 8**, three of the NDD/ID related controls had an IFN score slightly above the cut-off value of 2.466 defined by Rice et al. to be abnormal [49]. However, the IFN scores of all related and unrelated controls remain significantly lower than those of the four tested NDD/ID patients, thereby confirming that these *PSMC3* variants were associated with enhanced type I IFN signaling.

**Figure 8:**
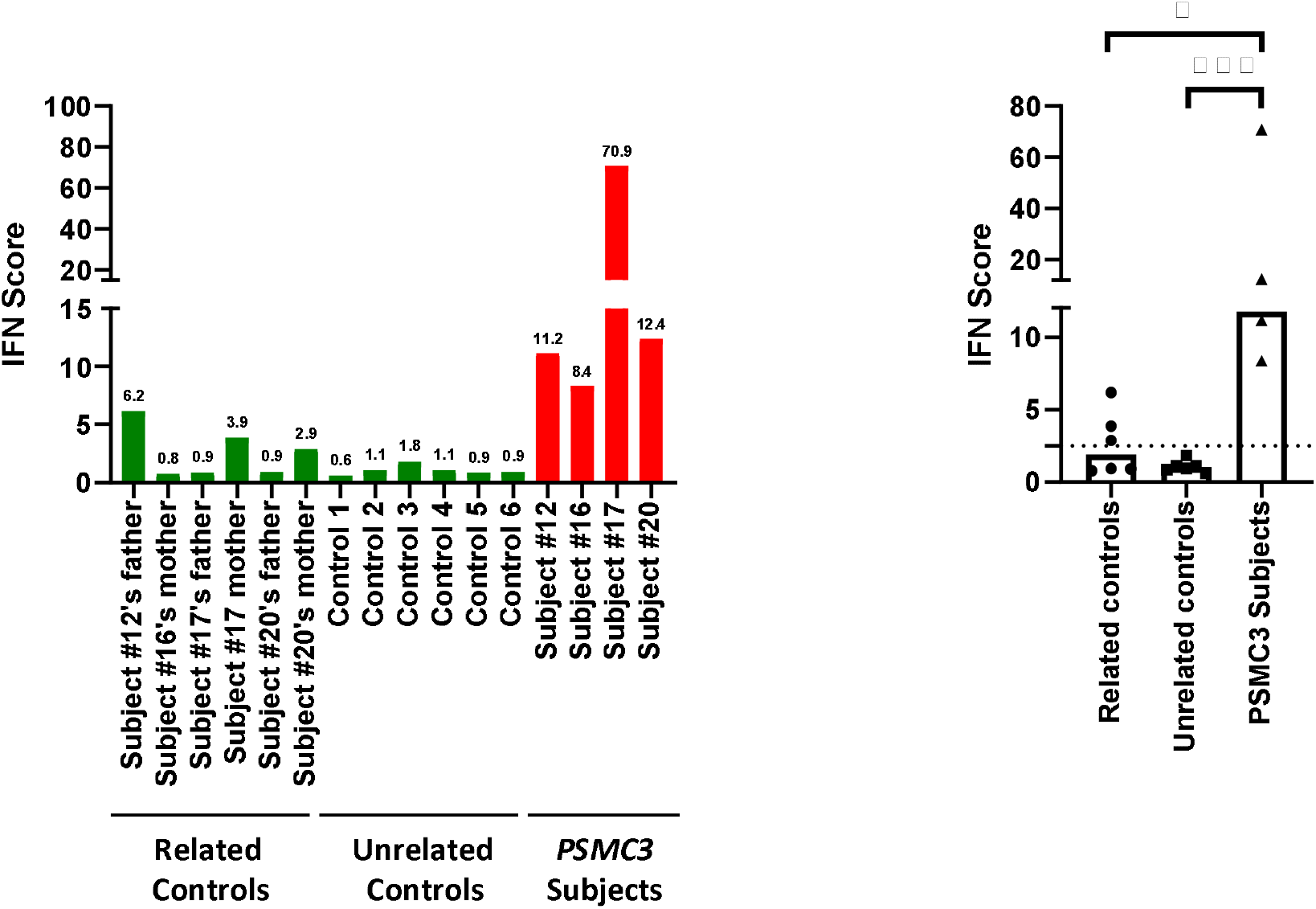
Interferon (IFN) score of *PSMC3* subjects suffering from NDD/ID. IFN scores for the NDD/ID families #12, #16, #17 and #20 (related controls and affected individuals) as well as for six unrelated controls (1 to 6) were calculated as the median of the RQ of the six ISG over a single calibrator control. Shown are the IFN scores of each sample (left panel) and the sample groups i.e. related controls, unrelated controls and *PSMC3* subjects, as indicated (right panel). Statistical significance was assessed by ratio paired t test where *indicates p<0.1 and ** indicates p<0.05.

## Discussion

Here, we report fourteen missense variants in the *PSMC3* gene in twenty-two unrelated individuals with NDD **(Figure 1, Tables 1 and 2)** which clearly identify the 19S AAA-ATPase proteasome subunit PSMC3/Rpt5 as a critical protein for the development of the central nervous system (CNS). This notion is in line with previous reports showing that conditional inactivation of other 19S proteasome subunits (i.e. PSMC1/Rpt2 and PSMC4/Rpt3) in mice results in severe neuronal phenotypes with features of neurodegeneration and locomotor dysfunction [50, 51].

Recently, a homozygous deep intronic variant creating a cryptic exon in the *PSMC3* gene was linked to a familial recessive neurosensory syndrome [52]. However, given their distinct modes of inheritance and pathogenesis, we believe that this recessive disorder observed in a single family and the dominant one, which we describe here in multiple families, are two different clinical entities. This assumption is substantiated by the fairly partial and inconstant overlap of disease phenotypes as neither cataract, nor neuropathy or similar cutaneous features –which are the hallmarks of the recessive syndrome- were observed in patients with NDD/ID reported in the present series. This notion is also supported by the fact that, unlike the familial recessive neurosensory syndrome described by Kroll et al., the dominant NDD/ID identified in this manuscript is not characterized by constitutive processing of TCF11/Nrf1 **(Figure 6B)**.

Cognitive flexibility is an important aspect of typical brain function which allows adaptation to both physical and social environmental changes [53, 54]. This may be assessed by evaluating reversal learning performance, a process which was identified several years ago in Drosophila adult flies [40] and whose dysfunction has been associated with the pathogenesis of various neuropsychiatric disorders [55–59]. Strikingly, while *PSMC3* gene-silencing in flies had no discernible effect on learning performance, it severely compromised reversal learning **(Figure 2)**. Interestingly, although proteasomes have been shown to regulate long-term potentiation (LTP) [60, 61], their involvement in reversal learning was not known. Our data clearly identify PSMC3/Rpt5 as a key regulator of this process whose molecular landscape was initially limited to a few molecules related to the cytoskeleton and GABAergic system [62–64]. Additional evidence in favour of a critical role of PSMC3/Rpt5 in behavioural flexibility emerges from our experiments in primary hippocampal neurons showing that PSMC3/Rpt5 overexpression affects dendrite growth **(Figure S4)**. One could argue that the adverse effects of PSMC3/Rpt5 on this process might be due to extra- proteasome functions as a consequence of an excess of “free” subunit following transfection. However, our investigations on patient T cells clearly show that *PSMC3* missense variants are associated with an increased accumulation of ubiquitin-modified proteins **(Figure 6A)**. This indicates that these alterations give rise to proteasome loss-of-function variants associated with perturbed protein homeostasis. Interestingly, our proteomic analysis revealed that subject T cells carrying *PSMC3* variants were particularly enriched with ribosomal proteins such as RPL4, RPL6, RPL7A and RPL7 **(Figure 5 and Table 3)**. These proteins may be specifically targeted for degradation in these affected individuals and that their accumulation occurs as a consequence of impaired intracellular protein clearance. Consistent with this notion, proteasome inhibition has been recently shown to result in the aggregation of ubiquitin-modified ribosomal proteins [65]. Our data therefore support the recent view that ribosome dysregulation defines a key feature of NDD/ID phenotypes [66, 67] and the concept of translational arrest upon proteotoxic stress via the action of eIF2a kinases **(Figure 5A)**.

**Table 3.** List of proteins enriched in *PSMC3* Subjects 16 and 20 versus their controls (mother and father/mother, respectively)

| **PG.Protein Groups** | **Protein name** | **Gene name** | **Mut/Co\|signal**<br>**_log2_ratio** | **Mut/Co\|raw_p_value** | **Mut/Co\|adjusted_p_value** |
| --- | --- | --- | --- | --- | --- |
| P02749 | Beta-2-glycoprotein 1 | APOH | 3,90875773 | 3,9233E-05 | 0,01752392 |
| P01861 | Immunoglobulin heavy constant gamma 4 | IGHG4 | 3,41081457 | 3,9313E-07 | 0,0003951 |
| Q02878 | 60S ribosomal protein L6 | RPL6 | 3,09519982 | 8,1085E-05 | 0,02173071 |
| P62424 | 60S ribosomal protein L7a | RPL7A | 2,97520531 | 2,4946E-06 | 0,00143263 |
| P18124 | 60S ribosomal protein L7 | RPL7 | 2,90834996 | 5,5264E-05 | 0,01815176 |
| P83731 | 60S ribosomal protein L24 | RPL24 | 2,78639617 | 5,8732E-05 | 0,01815176 |
| P16403 | Histone H1.2 | H1-2 | 2,72232694 | 4,4042E-05 | 0,0177049 |
| P02768 | Serum albumin | ALB | 2,64236248 | 3,3722E-18 | 1,3556E-14 |
| P36578 | 60S ribosomal protein L4 | RPL4 | 2,50022766 | 3,276E-07 | 0,0003951 |
| P01876 | Immunoglobulin heavy constant alpha 1 | IGHA1 | 2,32067402 | 3,007E-05 | 0,01511011 |
| P20591 | Interferon-induced GTP-binding protein Mx1 | MX1 | 2,20162951 | 6,3215E-05 | 0,01815176 |
| Q6P2Q9 | Pre-mRNA-processing-splicing factor 8 | PRPF8 | 1,33941179 | 1,5549E-06 | 0,00125012 |
| Q01469 | Fatty acid-binding protein 5 | FABP5 | -2,29088412 | 5,7962E-05 | 0,01815176 |

One key finding is the observation that NDD/ID subjects with *PSMC3* variants generate a type I IFN gene signature with elevated expression of typical ISG including *IFIT1*, *IFI27*, *IFI44*, *IFI44L*, *ISG15* and *RSAD2* **(Figures 7 and 8)**. Although it is well-established that proteasome loss-of-functions variants cause interferonopathies in CANDLE/PRAAS subjects, these alterations were long restricted to subunits of the 20S core particle and/or proteasome assembly factors so far [29–35]. It was only recently that pathogenic variants in subunits of the 19S regulatory particle, namely PSMD12, were reported to engage constitutive type I IFN signaling in patients with Stankiewicz-Isidor syndrome (STISS) (in press), a NDD disorder sharing levels of similarities with the one described in this manuscript. Intriguingly, although both CANDLE/PRAAS subjects and NDD/ID individuals with *PSMD12* or *PSMC3* variants carry genomic alterations that affect the same multi-subunit enzyme (*i.e*. 26S proteasome), their clinical phenotypes do not entirely overlap. Notably, NDD/ID subjects with *PSMC3* variants failed to develop recurrent fever, lipodystrophy and/or skin lesions which are usually detected in CANDLE/PRAAS subjects **(Table 1)**. One could argue that such differences may reflect distinct localizations of the affected subunits within the 26S proteasome complex, thereby suggesting that alterations of the 19S regulatory particle promote the generation of NDD/ID, while those of the 20S core particle and/or assembly chaperones favor the development a CANDLE/PRAAS phenotype. This assumption is however challenged by the fact that PSMB1/β6 variants of the 20S core particle lead to the acquisition of neuronal phenotype very similar to that seen in NDD/ID subjects with *PSMC3* variants [27]. The lack of systemic autoinflammation in NDD/ID subjects mounting a constitutive type I IFN response may seem surprising at first sight, but it is not totally unexpected, since this inconsistency is found in other neurodevelopmental disorders including Aicardi-Goutières [68, 69] and Down syndromes [70, 71]. This is particularly well exemplified in Down syndrome patients who, like NDD/ID subjects with *PSMC3* dominant variants, exhibit a constitutive activation of type I IFN signaling [72, 73]. To what extent type I IFN actively contributes to the pathogenesis of these disorders remains to be fully determined, even though a growing body of evidence supports the notion that IFN has detrimental effects on CNS function [74–77] and/or stem cell function and differentiation [78, 79].

Because proteasome dysfunction typically engages stress responses involving compensatory mechanisms such as autophagy [80], the integrated stress response (ISR) and the unfolded protein response (UPR) [26, 81], we reasoned that the sterile type I IFN response detected in subjects with *PSMC3* variants might be due to sustained activation of either one of these pathways. As anticipated, high levels of autophagy and ER stress were detected in NDD/ID affected individuals, as evidenced by increased expression of the LC3-II and GRP94 proteins **(Figure 6A)**. Both PINK1 and NIX mitochondrial proteins were found to be decreased in NDD/ID subjects **(Figure 6B)**, suggesting that *PSMC3* variants increase autophagy-driven elimination of mitochondria (i.e. mitophagy). This observation supports the growing consensus that mitochondria dysfunction is a key determinant of the pathogenesis of neurodevelopment [82]. Besides, the activation status of PKR, a protein of the ISR that intersects with the UPR [83], was substantially increased in all investigated *PSMC3* subjects. Both ISR and UPR have the ability to counterbalance proteotoxic stress by inducing a global translational arrest via eIF2a phosphorylation. This is accompanied by concomitant accumulation of non-translated mRNAs, the formation of stress granules recruiting different RNA species and RNA processing enzymes, and IRE-1 dependent mRNA decay (RIDD) [84]. Although PKR typically responds to viral double stranded RNA [85], it also may undergo activation under sterile conditions upon different stresses including ER-stress involving PKR-associated activator PACT and its modulator TRBP, a protein required for micro RNA biogenesis [86–88]. Both, PACT and TRBP were observed to be increased in subject’s cells along with a couple of RNA-processing factors in our omics analysis strongly indicating that PKR activation occurs via uncontrolled RNA processing. The mechanisms by which *PSMC3* variants activate PKR in NDD/ID affected individuals remain unclear, but our data open the possibility that PKR may sense a broader spectrum of danger signals than initially assumed, including perturbations of protein homeostasis. This concept is in line with the observation that activated PKR was found in the CNS of subjects with neurodegenerative diseases [89–92] and that neurodegeneration is associated with neuroinflammation [93, 94]. Altogether, our work demonstrates that heterozygous *PSMC3* dominant variants result in a neurodevelopmental syndrome associated with a specific type I IFN gene signature.

## Data Availability

All data produced in the present study are available upon reasonable request to the authors

## Acknowledgements

This work was supported by the German Research Foundation (SFBTR167, RGT2719-PRO project B4) to EK, E-Rare project GENOMIT (Austrian Science Fund FWF, I4695-B) to JAM, in part, by US National Institutes of Health (NIH) grants (R01MH101221) and a grant from the Simons Foundation (SFARI #608045) to E.E.E.; E.E.E. is an investigator of the Howard Hughes Medical Institute.

