## Supplements for "De novo variants in the *PSMC3* proteasome AAA-ATPase subunit gene cause neurodevelopmental disorders associated with type I interferonopathies"

**Supplemental Materials & Methods**

**Supplemental Figures S1, S2, S3, S4 and S5.**

**Supplemental Materials & Methods**

**Proteomic studies**

*Preparation of protein samples* -- Protein was extracted from primary T cells by five cycles of freezing (liquid nitrogen) and thawing (30^o^C, 1,400 rpm) in 8 M urea/ 2 M thiourea. Cell debris and insoluble material was separated by centrifugation (16,000 × g, 1 h at 20 C). Protein content was determined with a Bradford assay (Biorad, Munich, Germany).

*Sample preparation* *for mass spectrometry*-- Four µg of total protein from each sample were reduced (2.5 mM DTT ultrapure, Invitrogen, for 15 min at 37 °C) and alkylated (10 mM iodacetamide, Sigma Aldrich, for 30 min at 37°C). Benzonase (Novagen, Merck Millipore, Darmstadt Germany) 0.6 U/µg protein) was applied for nucleic acid degradation before digestion with trypsin (Promega, Madison, WI, USA) at an enzyme to protein ratio 1:25 over night at 37 °C. The tryptic digestion was stopped by adding acetic acid (final concentration 1%) followed by desalting using ZipTip-µC18 tips (Merck Millipore, Darmstadt, Germany). Eluted peptides were concentrated by evaporation under vacuum and subsequently resolved in 0.1% acetic acid, 2% acetonitrile (ACN) containing HRM/iRT peptides (Biognosys, Zurich, Switzerland) according to manufacturer’s recommendation. For the generation of a spectral library a protein pool of equal protein amounts (10 µg) of each sample was prepared as described above. Resulting peptides were desalted (SepPak, Waters, Eschborn, Germany) and fractionated by strong cation exchange chromatography (PolySulfoethyl A™ column, 150 x 1mm, 5µm, 200A, Poly LC Inc., Columbia, ML, USA). Peptide fractions (n=17) were purified for mass spectrometric analyses as described above.

*Mass Spectrometry Measurements* -- Mass spectrometric (MS) data was recorded on a QExactive HF mass spectrometer (Thermo Electron, Bremen, Germany). Before MS data acquisition tryptic peptides were separated by reverse phase chromatography (Accucore 150-C18, 25 cm x 75 μm, 2,6 μm C18, 150 Å) using an Ultimate 3000 nano-LC system (both Thermo Scientific, Waltham, MA, USA) at a constant temperature of 40°C and a flow rate of 300 nL/min. To design a spectral library, MS/MS peptides were separated by 120 min linear gradients with increasing acetonitrile concentration from 5 to 25 % in 0.1 % acetic acid. Data were recorded in data dependent mode (DDA). The acquisition of MS data for relative quantitation was performed in data independent mode (DIA) after peptide pre-fractionation at chromatographic conditions described above (see Table S2). For further details to the instrumental setup and the parameters for LC-MS/MS analysis in DDA and DIA mode.

*Data analysis* -- Proteins were identified using Spectronaut^TM^ Pulsar 13.9 software (Biognosys AG) against a spectral library generated from data-dependent acquisition measurements of a SCX-fractionated peptide pool. The spectral library construction by Spectronaut was based on a database search using a human protein database (Uniprot vs 03_2019, 20404 entries). The generation of the ion library in Spectronaut^TM^ v13.9.191106.43655 resulted in a constructed library consisting of 920,617 fragments, 67,323 peptides and 6717 protein groups. The Spectronaut DIA-MS analysis was carried out as described previously ^1^ with project specific modifications. Peptides were assigned to protein groups and protein inference was resolved by the automatic workflow implemented in Spectronaut. Only proteins with at least two identified peptides were considered for further analyses. Data analysis was performed with an in-house R-pipeline. Data was median normalized on ion level. Statistical analysis was carried out on peptide level using the algorithm ROPECA ^2^. Peptides with oxidized methionine were not included in the quantitative analysis. Binary differences have been identified by application of a moderate paired t-test ^3^. Family was used as pairing variable. Multiple test correction was performed according to Benjamini-Hochberg. Variance within the data set was visualized by principal component analyses and differences in the protein pattern by Volcano plots. For representation of protein intensities Hi3Peptides were used.

**Gene expression analysis by Nanostring**

One hundred nanograms of total RNA isolated from control and patients was hybridized to Nanostring nCounter® Human AutoImmune Profiling Panel and quantification of gene expression was subsequently performed following the manufacturer’s instruction following normalization to housekeeping genes.

**Facial image analysis**

We utilized GestaltMatcher ^4^ to measure the similarities of the facial phenotypes between individuals with mutations in PSMC3. This approach has been used to quantify the intra-syndromic similarity, as well as the inter-syndromic similarity, which can also reflect interaction on a molecular level ^5-7^. GestaltMatcher spanned a 320-dimensional clinical face phenotype space (CFPS) defined by the network parameters of DeepGestalt ^8^. DeepGestalt itself is a deep convolutional neural networks (DCNN) that was trained on 20,091 frontal images of 299 different syndromes. For GestaltMatcher, we encoded each image first by a 320-dimensional facial phenotypic descriptor, which is equivalent to its position in the CFPS. The similarity between two images was quantified by the cosine distance. In CFPS, the images with close distance were considered to have a high overlap of syndromic facial features. Therefore, we ranked the patients by sorting the cosine distance. We performed pairwise comparisons on the 13 photos of 12 subjects (S-2, S-4, S-5, S-6, S-9, S-11, S-12, S-15, S-16, S-17, S-20, and S-22) and together with 3,533 images from 2,516 diagnosed patients with 816 syndromes in Face2Gene database. There were two photos for S-22 that were taken at different ages. The following two criteria selected the 2,516 patients from the database. A patient's diagnosed syndrome was not included in the model training and had less than seven subjects. By these selection criteria, we formed the CFPS with syndromes that have not been seen by the model and have very few subjects simulating the ultra-rare diseases.

**Neuronal morphology assays**

PSMC3 genes (WT and mutated) were placed each in our dual promotor expression vector ^9^, to be able to test them in the in vitro neuronal morphology assay. For neuronal cultures, FvB/NHsD females were crossed with FvB/NHsD males (both ordered at 8-10 weeks old from Envigo). All mice were kept group-housed in IVC cages (Sealsafe 1145T, Tecniplast) with bedding material (Lignocel BK 8/15 from Rettenmayer) on a 12/12 h light/dark cycle in 21°C (±1°C), humidity at 40-70% and with food pellets (801727CRM(P) from Special Dietary Service) and water available ad libitum. All animal experiments were conducted in accordance with the European Commission Council Directive 2010/63/EU (CCD approval AVD101002017893).

Primary hippocampal neuronal cultures were prepared from FvB/NHsD wild-type mice according to (Goslin and Banker 1991). Briefly, hippocampi were isolated from brains of E16.5 embryos and collected altogether in 10 ml of neurobasal medium (NB, Gibco) on ice. The samples were incubated in pre-warmed trypsin/EDTA solution (Invitrogen) at 37°C for 20 minutes, and dissociated using a 5 ml pipette in 1.5 ml NB medium supplemented with 2% B27, 1% penicillin/streptomycin and 1% glutamax (Invitrogen). Following dissociation, neurons were plated in a small drop on poly-D-lysine (25 mg/ml, Sigma) coated 15 mm glass coverslips at a density of 1*106 cells per coverslip in 12 well plates containing 1 ml of supplemented NB for each coverslip. The plates were stored at 37°C/5% CO2 until the day of the transfection. Neurons were transfected after 3 days in vitro (DIV) with the following DNA constructs: empty vector (1.8 µg per coverslip), PSMC3WT, PSMC3Met175Val, PSMC3Arg304Trp, PSMC3Glu305Asp or PSMC3Glu383Lys (all 2.5 µg per coverslip). Lipofectamine 2000 was used to transfect neurons, according to the manufacturer’s instructions (Invitrogen). For the neuronal morphology analysis, neurons were fixed 5 days post-transfection with 4% paraformaldehyde (PFA)/10% sucrose, and incubated overnight at 4°C with MAP2 (1:500, #188004, Synaptic System) in GDB buffer (0.2% BSA, 0.8 M NaCl, 0.5% Triton X-100, 30mM phosphate buffer, pH7.4). Next day the neurons were incubated for 1 h in the anti-guinea-pig-Alexa647 (#706-605-148) conjugated secondary antibody (1:200, Jackson ImmunoResearch). Slides were mounted using Mowiol-DABCO (Sigma) mounting medium. Confocal images were acquired using a LSM700 confocal microscope (Zeiss).

For the analysis of the neuronal transfections, at least ten confocal images (20X objective, 0.5 zoom, 1024x1024 pixels) of different transfected neurons (identified by the red staining from the tdTomato) were taken from each coverslip for each experiment with at least two independent replications. For the analysis of the neuronal morphology, the NeuronJ plugin for ImageJ software was used to trace the dendrites with their branches. Total neurite length and arborization were measured and analyzed. All values were normalized against the mean value for each parameter of the control (control vector). Analysis was done by an experimenter blinded for the transfection conditions.

**Data representation and statistical analyses**

Neuronal morphology data was analyzed using a One-Way ANOVA, followed by a Tukey’s post-hoc test for multiple comparisons. All charts and statistical analyses were generated using GraphPad version Prism 8. A p value <0.05 was considered significant. All data are available on request from authors.

**Supplemental figures**

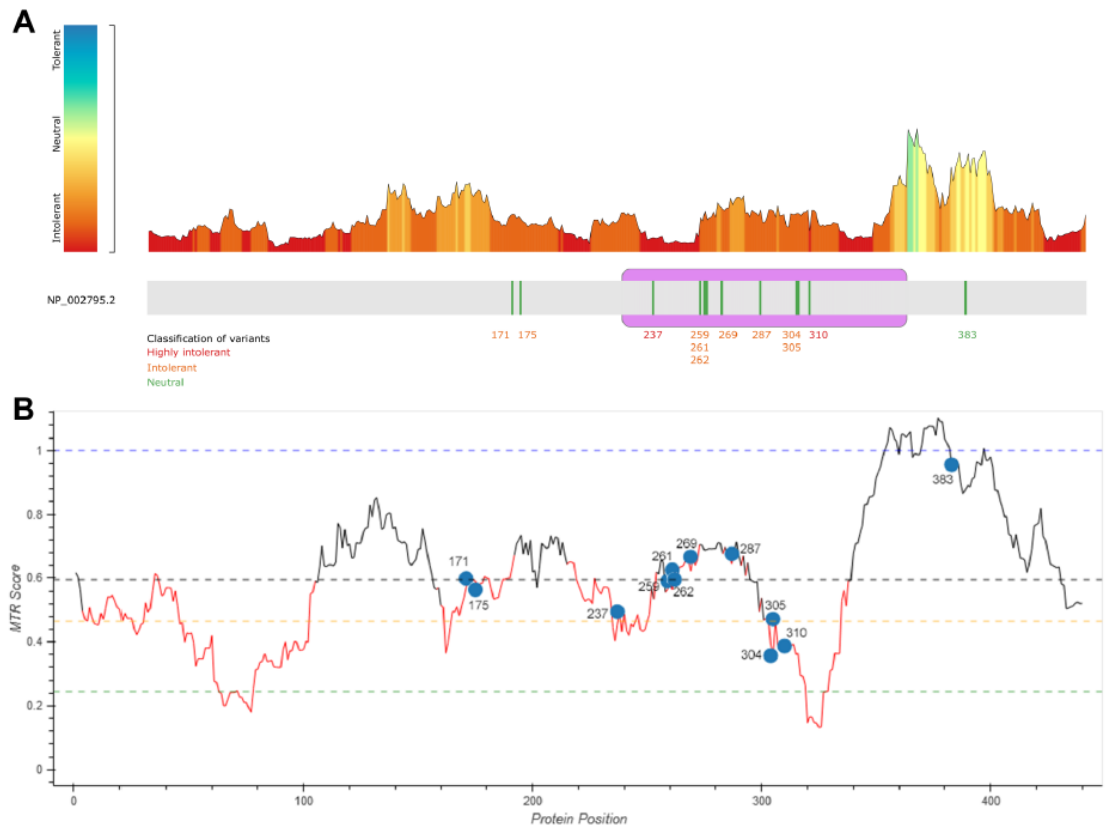

**Figure S1. Tolerance predictions of the amino acid residues of PSMC3/Rpt5 (NP_002795.2) affected by the variants reported in the study.** *A*. MetaDome web tool (PMID: 31116477) points to the general intolerance of *PSMC3* to missense variants. In consequence, all variants save p.(Glu383Lys) affect residues predicted intolerant or highly intolerant to variations. *B*. Analysis by Missense Tolerance Ratio (MTR; v1) confirms this trend and suggests that the variants reported affect intolerant residues and are expected to bear a severe effect, by contrast to p.(Glu383Lys) which impact on *PSMC3* function is more uncertain. Horizontal lines show gene-specific MTR percentiles 5th (in green), 25th (in yellow), 50th (in black), and neutrality (in blue; MTR = 1.0) MTR calculated using WES component of gnomAD v2.0.

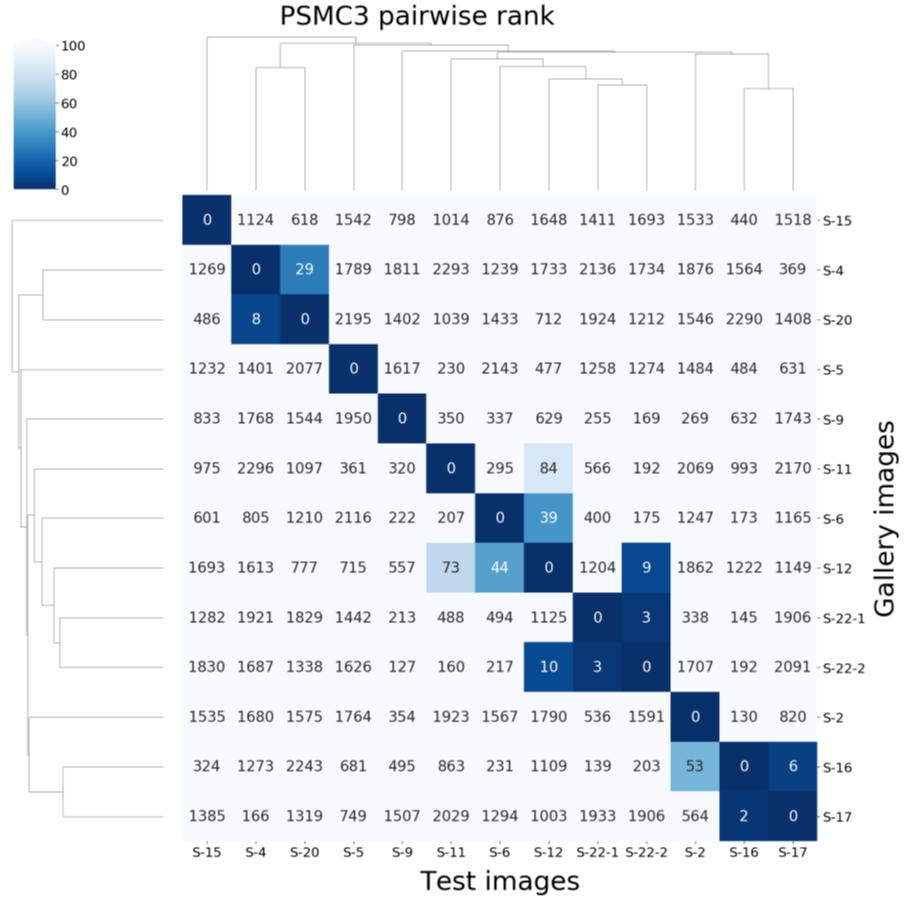

**Figure S2. Pairwise rank and hierarchical clustering of 13 photos in CFPS.** Gallery images were the images in CFPS which can be matched. Each column is the result of testing one subject in the column and listing the rank of the rest ten photos in each row. For example, by testing S-16, S-17 was on the 2nd rank and S-16 was on the 6th rank of S-17.

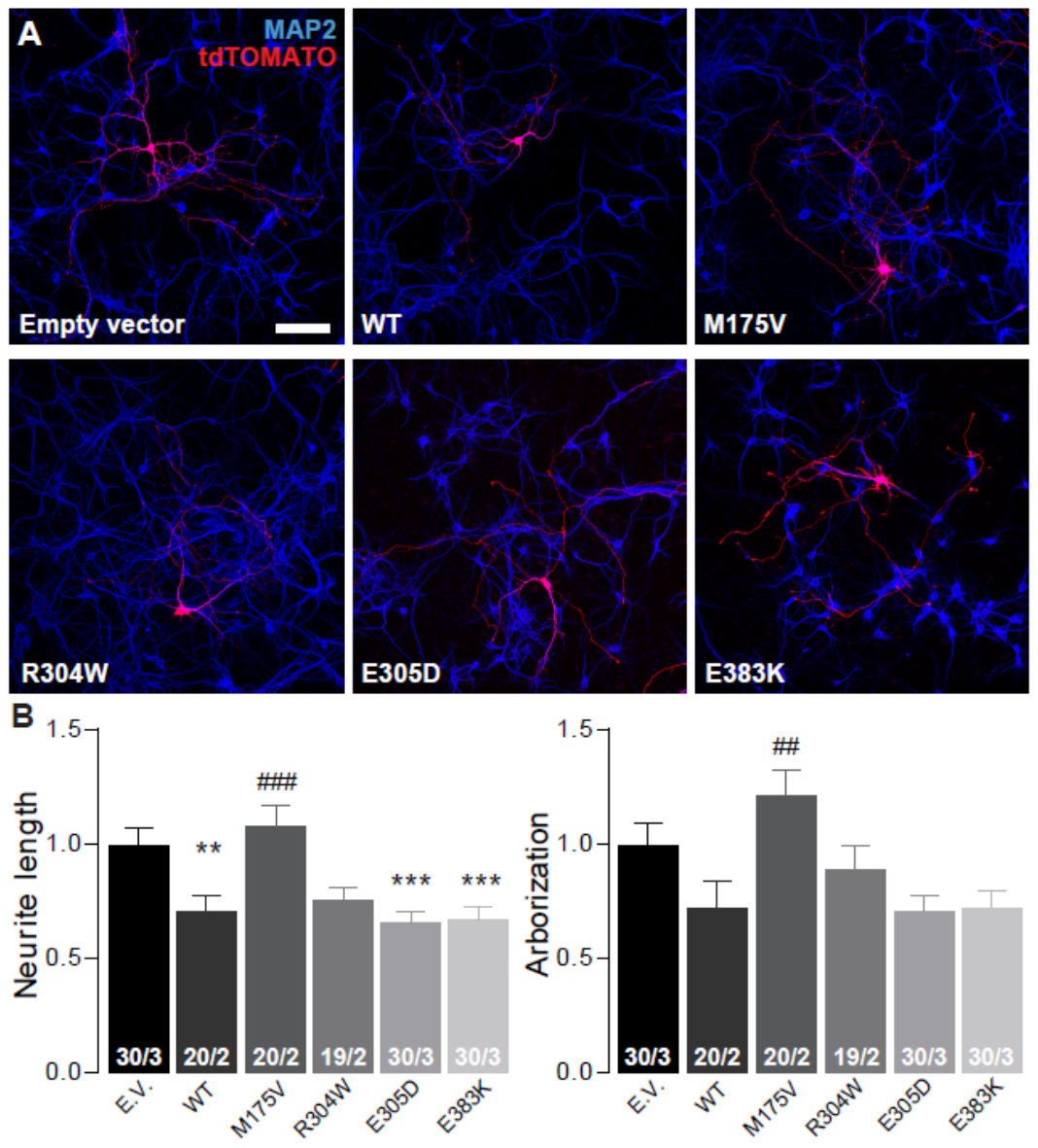

**Figure S3. Overexpression of PSMC3 is damaging for neuronal development in vitro**. *A*. Representative confocal images of primary hippocampal neurons transfected with empty vector (EV), PSMC3WT, PSMC3M175V, PSMC3R304W, PSMC3E305D or PSMC3E383K, where the tdTOMATO positive neurons express the different conditions. Scale bar 100 µm. *B*. Quantification of the neuronal morphology with total neurite length and arborization normalized to the empty vector control. Data are presented as mean ± SEM. Number of independently analyzed culture wells/plugs is indicated for each condition. ** p<0.01, *** p<0.001 compared to empty vector; ## p<0.01, ### p<0.001 compared to PSMC3WT.

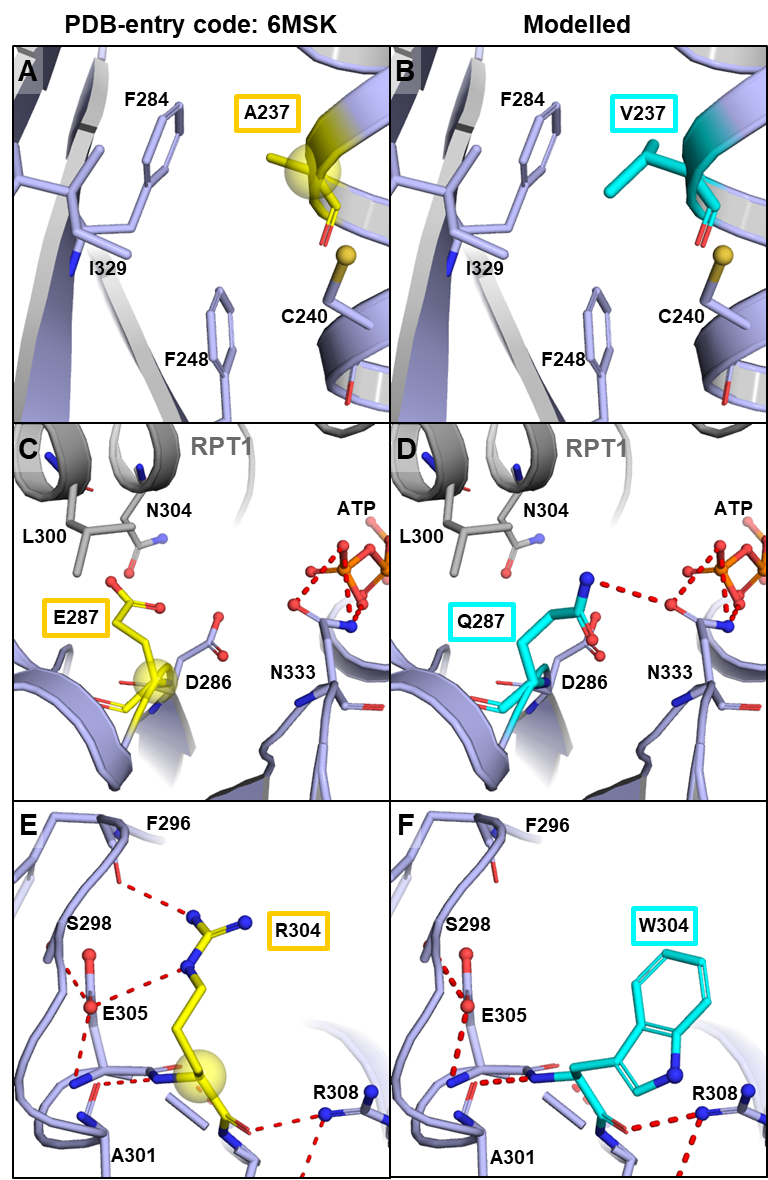

**Figure S4. Detailed representation of the *PSMC3* missense variants p.(Ala237Val), p.(Glu287Gln) and p.(Arg304Trp).** Substitution of alanine (*A*) into valine (*B*) at position 237 within PSMC3/Rpt5 is difficult to classify on a structural level. It can only be assumed that the increased size of Val237 compared to Ala237 disrupts the optimal packing of sidechains in the area. Glu287 is in close proximity to the ATP binding site of PSMC3/Rpt5 (*C*). Substitution into Gln287 (*D*) is likely to create additional polar bonds with N333 and might influence ATP binding and/or hydrolysis. Substitution of the positively charged Arg304 (*E*) into a bulky aromatic Trp304 (*F*) presumably interrupts the polar network and leads to steric clashes. To illustrate the differences between wild-type amino acid and the respective mutation, both are juxtaposed in the structural context of the wild-type one (left wild type, right mutation), although it can be assumed that the overall structure adapts to the mutation and thus structural changes not recorded here occur.

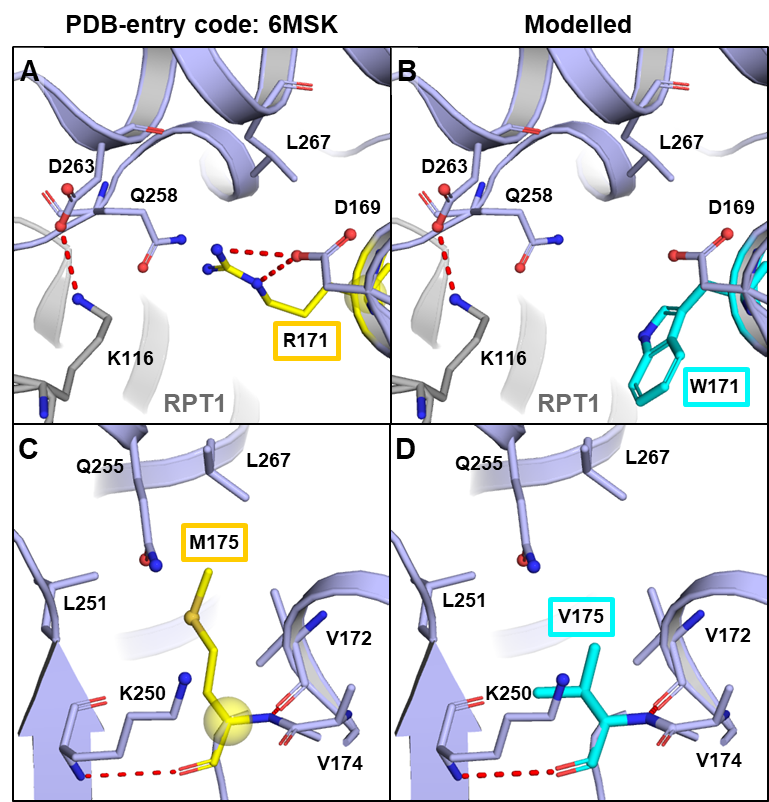

**Figure S5. Detailed representation of the *PSMC3* missense variants p.(Arg171Trp) and p.(Met175V).** Arg171 is located at the PSMC3/Rpt5(blue)-PSMC2/Rpt1(grey) interface (A) and forms polar interactions with Asp169 and Asn258. Substitution of the positively charged Arg171 (*A*) into a bulky aromatic Trp171 (*B*) presumably interrupts the polar network and leads to steric clashes in the 6MSK structure. Mutation of Met175 (C) to Val175 (D) is associated with the loss of a polarized sulphide group and therefore expected to diminish the hydrophilic environment required for the stabilisation of tertiary structures. Mutational sites in the wildtype are shown in yellow the C-α atom is indicated as yellow sphere. Variants are coloured turquoise. To illustrate the differences between wild-type amino acid and the respective mutation, both are juxtaposed in the structural context of the wild-type one (left wild type, right mutation), although it can be assumed that the overall structure adapts to the mutation and thus structural changes not recorded here occur.
